# Personalized planning of cardiac resynchronization therapy through integration of coronary sinus geometry, clinical data, digital twins, and machine learning: visualization, stratification, and optimization

**DOI:** 10.64898/2026.07.01.26356827

**Authors:** Anastasia Bazhutina, Tatiana Chumarnaya, Stepan Zubarev, Margarita Budanova, Vera Stepanova, Svyatoslav Khamzin, Dmitry Lebedev, Olga Solovyova

## Abstract

**Background:** Cardiac resynchronization therapy (CRT) fails in 30% of patients, often due to suboptimal left ventricular pacing site (LVPS) selection. Current practice lacks tools for pre-procedural, patient-specific LVPS optimization within the accessible coronary sinus (CS) tributaries. This study aimed to develop a digital twin and an explainable ML-based clinical decision support framework to address this issue.

**Methods:** Personalized 3D cardiac models incorporating ventricular anatomy, myocardial fibrosis, and CS anatomy were constructed from CT and LGE-MRI for 74 CRT candidates. Finite-element Eikonal simulations of biventricular pacing generated patient-specific electrophysiological features at candidate LVPS. A Machine Learning (ML) classifier was trained on a hybrid feature set of pre-procedural clinical variables and model-derived indices, validated by leave-one-out cross-validation. SHAP analysis provided a physiologically interpretable rationale for each prediction. The framework was applied to a pilot cohort of 19 patients with reconstructed 3D CS anatomy to generate a spatial likelihood map of CRT response across all clinically implantable pacing sites within each patient’s CS.

**Results:** The ML classifier outperformed the reference Feeny clinical calculator under LOO-CV (accuracy 0.78 vs 0.58; F1-score 0.75 vs 0.43), AUC=0.78, sensitivity=0.80, specificity=0.77. Bootstrap analysis yielded mean AUC=0.85 (95% CI 0.70–0.95). In the pilot CS cohort, the framework identified that 8 of 13 clinical non-responders had no accessible CS site predicted to yield a positive response, supporting redirection towards alternative pacing strategies. In the remaining 5, alternative implantable sites with high predicted response probability were identified. SHAP analysis confirmed that dominant predictors were patient-specific in their relative contributions, supporting individualized over heuristic-based LVPS selection.

**Conclusion:** This pilot study demonstrates the feasibility of a digital twin and explainable ML framework as a pre-procedural clinical decision support tool for CRT planning, stratifying patients and identifying optimal implantable sites with transparent anatomical rationale. Prospective validation and regulatory evaluation are required before clinical deployment.

## 1 Introduction

Cardiac resynchronization therapy (CRT) is an established treatment for patients with chronic heart failure and baseline electrical dyssynchrony, with a proven reduction in mortality, hospitalizations, and symptom burden. However, despite carefully applied guideline-based selection, the procedure fails to produce a clinically meaningful response in 30% of implanted patients. As a result, CRT has one of the highest non-response rates among recommended cardiac device therapies [1]. According to current guidelines, a conventional CRT configuration for biventricular pacing (BiVP) with a transvenous left ventricular (LV) pacing lead implantation remains the first-choice procedure in a majority of CRT candidates [2, 3]. Therefore, a selection of pacing sites, especially the left ventricular pacing site (LVPS) within the accessible tributaries of the coronary sinus (CS) directly determines the pattern of ventricular resynchronization and a long-term effect of pacing. Yet, the selection is currently made intraoperatively without pre-procedural, patient-specific guidance.

Numerous strategies have been proposed to guide LVPS selection, including targeting the latest activated area (LAA) of the LV [4, 5], avoiding myocardial fibrosis [3, 6], and maximising the QLV sensing delay [7, 8]. Non-invasive electroanatomical mapping [9, 10] and echocardiographic assessment of mechanical dyssynchrony [11] have also been explored to define optimal target veins. Recent artificial intelligence (AI) approaches have further demonstrated the ability to improve patient stratification for CRT response prediction from pre-procedural data [12]. However, three fundamental limitations remain across this body of work. First, no approach constrains a multivariable optimization to the patient-specific CS anatomy, myocardial substrate, and the electrophysiological consequences of pacing at a specific candidate LVPS. Second, as noted in a recent systematic review [12], none of the reviewed AI models links response prediction directly to LVPS location within the accessible CS anatomy. Third, despite significant evidence accumulation, no LVPS optimization strategy has been adopted into current guidelines as a standard recommendation [2], reflecting the absence of a robust, multivariable pre-procedural planning tool. For patients in whom no suitable CS site can be identified, alternative approaches including left bundle branch area pacing and conduction system pacing are emerging options, further motivating the development of pre-procedural tools that can stratify such patients before implantation.

Personalized cardiac digital twins are patient-specific computational models that integrate individual anatomy, myocardial substrate, and electrophysiology. Digital twins provide a systematic approach to solving this multivariable planning problem [13–16]. A few studies have applied digital twin approaches to CRT optimization, combining personalized electrophysiological and electromechanical models with AI-based prediction [17–19]. In our previous work [18], we demonstrated that a supervised machine learning (ML) classifier trained on combined clinical and simulation-derived features can predict LVPS dependent CRT response likelihood at the individual patient level. However, this approach evaluated pacing sites across the entire LV epicardial surface without considering the anatomical constraints of the CS tree as the space where conventional implantation of the LV lead practically occurs [16, 20–22].

In this work, we set the following goals to overcome several limitations. The first goal was to extend the framework [18] to integrate patient-specific ventricular and CS digital twin models for a cohort of CRT patients, which allowed us to constrain LVPS optimization to accessible locations for implantation procedure. The second was to adjust the platform for patients with extensive fibrosis based on the detailed MRI-based fibrosis mapping and CT-derived CS anatomy. The latter data was unavailable in the patient cohort used in [18]. Therefore, combination of the dataset from [18] with an additional dataset collected for this work required adaptation of the ML pipeline with adapted preprocessing and nested hyperparameter optimization.

Here, we present a new computational framework based on digital twins and ML providing a 3D visualization of the spatial map of ML-scores predicting CRT response across implantable CS LVPS locations backed up by SHAP-based explanations. The approach has great potential to support patient-level stratification and implantation targeting.

These outputs are intended to support pre-procedural planning by translating personalized simulation data into spatially specific, interpretable guidance for clinicians.

## 2 Methods

### 2.1 Study Design and Participants

This retrospective study included clinical data from 74 CRT candidates evaluated at Almazov National Medical Research Centre. The study was approved by the institutional Ethics Committee, and written informed consent was obtained from all participants. Patients were selected according to current CRT guidelines, presenting with NYHA HF functional classes II–III, Left Ventricular Ejection Fraction (LVEF) *<* 35%, and left bundle branch block (LBBB) pattern according to the Strauss criteria [23] with QRS duration *>* 120 ms. The full statistics for the total group (n = 74) are described in Table S1 in the Supplementary.

Two cohorts were combined for classifier development. The **Non-CS group** (n=55) comprised data collected by us earlier from patients with standard clinical, ECG, echocardiographic, CT, and LGE-MRI data, with simplified AHA segment-based fibrosis modelling, and without CS geometry (see [18] for details). **The CS group** (n=19) comprised data from patients with dedicated contrast-enhanced CT venography enabling 3D CS anatomy reconstruction, and detailed LGE-MRI-based fibrosis modelling (Table 1). Clinical and imaging data were anonymized prior to analysis. In our study, each patient from the Non-CS group and CS group underwent standard pre-implant evaluation and follow-up in 12 months after the procedure, including 12-lead ECG and echocardiography. Positive response to CRT was defined using echocardiography as LVEF improvement greater than 10% at the follow-up as compared with baseline similar to [24]. The entire cohort (n = 74) included 30 responders (40.5%) and 44 non-responders (59.5%).

**Table 1.** Summary of the clinical, imaging, and model-derived indices in the CS patient group. Data is shown as a Mean±SD for normal distribution, a Median [25th percentile; 75th percentile] for non-normal distribution. * - p*<*0.05, ** - p*<*0.01 LBBB vs CRT or LBBB vs BiVP. Comparisons between two dependent groups were made using Wilcoxon’s test for quantitative data and McNemar’s test for qualitative data. # - p*<*0.05, ## - p*<*0.01 Responders vs Non-responders. Comparison between two independent groups was carried out using the Mann-Whitney test for quantitative data and Pearson’s chi-square test for qualitative data. BMI – body mass index; AF – atrial fibrillation; QRSd – mean QRS duration in 12-lead ECG; EF - left ventricular ejection fraction measured baseline LBBB, EDV – end-diastolic volume of left ventricle measured baseline LBBB; ESV – end-systolic volume of left ventricle measured baseline LBBB; EDD – left ventricular end-diastolic diameter measured baseline LBBB; ESD – left ventricular end-systolic diameter measured baseline LBBB; FC CHF – functional class of congestive heart failure; Scar-LVPS Distance – a distance from the LVPS to the scar/fibrosis area; RVPS-LVPS Distance – distance between right and left ventricular pacing sites; LAA-LVPS Distance – distance from LV pacing site to latest activated area; MTV – myocardial tissue volume; LVTV – left ventricular myocardial tissue volume; ScarV –scar/fibrosis tissue volume; ScarV/LVTV – percentage of fibrosis tissue relative to LVTV; TAT95 -95% of total ventricular activation time; QRSd – QRS complex duration measured baseline LBBB; AD*_RV LV_* – difference in the total LV and RV activation time; AD*_ST LV_* – relative difference between mean activation time of LV free wall and septum; ML-score – output of the SVM-classifier of CRT response defined as more than 10% LV EF improvement.

|  | CS patient group n=19 |  |  |  |  |  |
| --- | --- | --- | --- | --- | --- | --- |
| Variable | Responders n=6 (32%) |  |  | Non-responders n=13 (68%) |  |  |
| Gender male/female | 5/1 |  |  | 10/3 |  |  |
| Age, year | 65 ± 6 |  |  | 66 ± 9 |  |  |
| BMI | 28.6 ± 8.5 |  |  | 27.2 ± 2.6 |  |  |
| History of AF | 0 (0%) |  |  | 1 (7.7%) |  |  |
|  | LBBB | CRT | Δ,% | LBBB | CRT | Δ,% |
| QRSd, ms | 192 ± 20 | 152 ± 12** | -20 ± 10 | 177 ± 14 | 148 ± 21** | -17 ± 1 |
| EF, % | 22.8 ± 5.3 | 40.8 ± 9.7** | 18.0 ± 8.6 | 27.0 ± 4.1 | 33.7 ± 3.7** | 6.7 ± 1.6# |
| EDV, ml | 211[183;373] | 178[151;254]* | -24[-32;-12] | 226[200;265] | 194[173;231]* | -9[-17;-8] |
| ESV, ml | 168[148;280] | 107[88;144]* | -43[-51;-27] | 159[136;196] | 128[108;158]* | -19[-21;-8]## |
| EDD, mm | 69 ± 8 | 62 ± 7 | -9 ± 8 | 70 ± 7 | 64 ± 8 | -8 ± 5 |
| ESD, mm | 60 ± 8 | 50 ± 8 | -16 ± 11 | 60 ± 8 | 54 ± 9 | -9 ± 7 |
| FC CHF : |  |  |  |  |  |  |
| I | 0 (0%) | 3 (42.9%) |  | 0 (0%) | 4 (30.8%) |  |
| II | 3 (42.9%) | 4 (57.1%) |  | 5 (38.5%) | 9 (69.2%) |  |
| III | 4 (57.1%) | 0 (0%) |  | 8 (61.5%) | 0 (0%) |  |
|  | CT/MRI derived data |  |  |  |  |  |
| Scar-LVPS distance,mm | 19[2;61] |  |  | 2[0;29] |  |  |
| RVPS-LVPS distance,mm | 110[105;124] |  |  | 100[91;127] |  |  |
| LAA-LVPS distance,mm | 51 ± 24 |  |  | 47 ± 25 |  |  |
| MTV,ml | 274.54 ± 123.51 |  |  | 224.44 ± 51.51 |  |  |
| LVTV,ml | 202.25 ± 76.26 |  |  | 166.78 ± 45.8 |  |  |
| ScarV, ml | 28 ± 19 |  |  | 47 ± 25 |  |  |
| ScarV/LVTV, % | 17 ± 12 |  |  | 28±15 |  |  |
|  | Model-derived data |  |  |  |  |  |
|  | LBBB | BiVP | Δ,% | LBBB | BiVP | Δ,% |
| TAT95, ms | 243 ± 90 | 117 ± 13* | -48 ± 16 | 199 ± 50 | 115 ± 21** | -40 ± 14 |
| QRSd, ms | 193 ± 25 | 149 ± 11** | -22 ± 9 | 186 ± 13 | 148 ± 21** | -21 ± 12 |
| AD<sub>RV LV</sub> , ms | 122 ± 9 | 11 ± 21** | -91 ± 18 | 103 ± 34 | 10 ± 23** | -91 ± 23 |
| AD<sub>STLV</sub> | 0.38 ± 0.07 | 0.11 ± 0.07** | -28 ± 13 | 0.34 ± 0.07 | 0.11 ± 0.06** | -23 ± 9 |
|  | CRT response prediction |  |  |  |  |  |
| ML-score | 0.64[0.26;0.80] |  |  | 0.30[0.25;0.35]# |  |  |

### 2.2 Overview of the Clinical Decision Support Workflow

The proposed framework was designed as a foundation for development of a pre-procedural tool for a clinical decision support system (CDSS), providing the implanting operator with patient-specific guidance before CRT lead placement. The intended application provides the pre-procedural planning review with AI-supported predictions on the potential success of BiVP and 3D visualization of achievable LVPS locations in the CS tributaries classified as yielding a positive response, where the CDSS output is made available alongside standard clinical and imaging data. The workflow proceeds in four sequential stages (Figure 1):

1. First, data collection and the construction of a digital twin. Pre-procedural CT (for cardiac and CS anatomy) and LGE-MRI (for fibrosis characterization) are processed to construct a patient-specific 3D cardiac finite-element model. The 12-lead ECG provides the electrophysiological calibration target for model parameter tailoring. Such data is already collected as a part of the routine pre-procedural workup at some centers performing advanced multimodality cardiac imaging, though it is not yet part of routine practice at most implanting centers.
2. Personalized finite-element models are then used to simulate the baseline (LBBB) ventricular activation and BiVP at multiple candidate LVPS across the clinically accessible CS tributaries. For each candidate LVPS, a set of anatomy-dependent indices and electrical dyssynchrony features is extracted.
3. The trained ML classifier is applied to the feature vector for each candidate LVPS, generating a continuous ML-score (0–1) representing the predicted probability of CRT response. Scores are mapped onto the 3D CS anatomy, producing a patient-specific spatial likelihood map of CRT response across accessible CS LVPS locations.
4. Finally, three CDSS outputs are delivered to the clinician: The patient-stratification verdict is indicative of the positive or negative prediction of response to conventional BiVP. The 3D likelihood map visualizes predicted ML-scores of the CRT response projected on a real CS anatomy. The optimal accessible LVPS of the highest ML score is identified and accompanied by a SHAP-based explanation that ranks the anatomical and electrophysiological drivers of that specific recommendation.

**Fig. 1.**
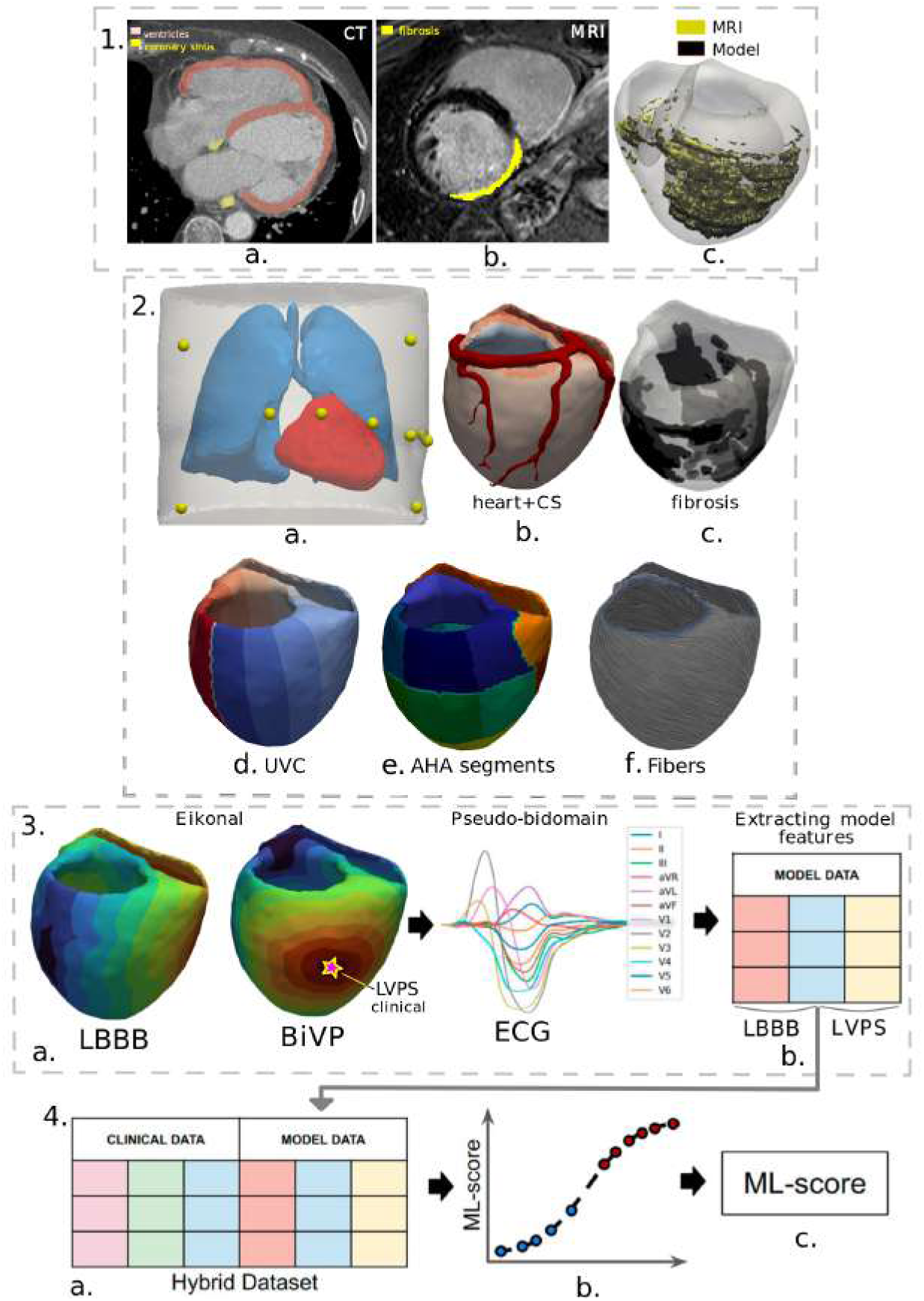
Digital twin construction and ML classifier development. (1,a–c) Multimodal segmentation from CT and LGE-MRI: torso, ventricles, lungs, myocardial fibrosis, and CS anatomy. (2,d–f) Finite-element model personalisation: UVC parametrisation, AHA 17-segment assignment, fibre orientation, and fibrosis integration. (3,a-b) Patient-specific simulations of LBBB activation and BiVP at the clinical LVPS; extraction of model-derived electrophysiological features. (4,a-c) Hybrid dataset construction by fusion of clinical and simulation-derived features; ML classifier training and ML-score generation.

The complete pipeline requires less than one hour on consumer-grade hardware from segmentation input to CDSS output. The framework does not autonomously determine clinical decisions; the physician retains full authority over the final implantation strategy. The CDSS output is designed to be reviewed at the planning meeting and documented in the pre-procedural record.

### 2.3 Core Computational Components

#### 2.3.1 Personalised Electrophysiology Modelling

Patient-specific 3D cardiac models were constructed from pre-procedural imaging data to enable personalised electrophysiological simulation of both baseline LBBB activation and BiVP. The pipeline for model generation consisted of the following key steps (Figure 1). Segmentation of the left and right ventricles, lungs, torso, and CS was performed manually from CT data; myocardial fibrosis was manually segmented from LGE-MRI and merged with the CT-derived ventricular anatomy. Tetrahedral volumetric meshes were generated using GMSH [25]. Universal Ventricular Coordinates [26] and AHA 17-segment labels were assigned using the approach described in [27]; myocardial fiber orientations were assigned using a rule-based method [28]. The propagation of electrical activation was simulated using an Eikonal model [29, 30], with anisotropic conduction properties (longitudinal-to-transverse conductivity ratio 4:1) and fibrosis regions assigned conductivity scaled by 0.3 relative to undamaged myocardium. Transmembrane action potentials were computed using the Ten-Tusscher Panfilov ionic model [31]; 12-lead ECGs were derived via pseudo-bidomain approach [32]. The models were calibrated electrophysiologically by fitting the simulated QRS duration to the patient’s clinical ECG under LBBB and BiVP conditions (see Figure S1 in the Supplementary) using the L-BFGS-B algorithm [33]. This personalization procedure yielded a cohort of 74 patient-specific finite-element models integrating individual anatomy, fibrosis geometry, and patient-specific electrical activation patterns.

#### 2.3.2 Hybrid dataset construction for CRT outcome prediction

A supervised ML classifier was developed to predict the probability of CRT response as a function of candidate LVPS location. The classifier was trained on a hybrid feature set integrating three categories of pre-procedural indices: (1) clinical and echocardiographic parameters; (2) CT/MRI-derived geometry-associated metrics, and (3) simulation-derived electrophysiological indices. The first category of indices included left ventricular ejection fraction (LVEF), end-systolic volume (ESV), end-diastolic volume (EDV), QRS duration (QRSd), and other standard clinical measurements.

The second category included indices derived from patient-specific biventricular finite element models reconstructed from CT data with fibrosis information. We computed three key geodesic distances by solving the isotropic Eikonal equation with the LV pacing site as the activation source. These distances were the time of the wavefront arrival from the LVPS to the nearest fibrosis region (Scar-LVPS), the time of arrival from the LVPS to late-activated myocardial regions (LAA-LVPS) at baseline LBBB activation, and the delay of wavefront arrival between the right ventricular (RV) and LV pacing sites (RVPS-LVPS). Additionally, we calculated volumetric parameters, including total myocardial tissue volume (MTV), left ventricular tissue volume (LVTV), scar/fibrosis volume (ScarV), and scar burden, expressed as a percentage of ScarV/LVTV. Detailed descriptions of these indices are provided in the Table 1 and the complete list of indices is described in Supplementary, Table S1.

The third category consisted of indices derived from personalised cardiac electrophysiology models and synthetic ECGs under conditions of both intrinsic LBBB activation and BiVP. These included temporal parameters: total activation time for 95% of the myocardium (TAT95) and model QRS duration (QRSd); interventricular electrical dyssynchrony index (activation delay between left and right ventricles *AD_RV LV_*), and intraventricular dyssynchrony measures *AD_ST LV_* and *IntAV_ST LV_*, which quantify activation differences between the LV lateral wall and septum; normalized indices - volume-normalized parameters *X_norm_*= *X/MTV*, where *X* is TAT95, QRSd, etc.; relative changes - delta values calculated as Δ*X* = (*X_BiV P_* − *X_LBBB_*)*/X_LBBB_*, where *X_LBBB_* and *X_BiV P_* represent index values during intrinsic LBBB activation and BiVP, respectively. These simulated metrics characterized the electrophysiological effects of BiVP and were subsequently used to optimize LV pacing site selection within the CS anatomy.

Features in categories (1) and the volumetric parameters (2) are patient-level constants. Geodesic distances and all BiVP-derived indices are recomputed for each candidate LVPS, enabling site-specific response prediction.

The binary outcome defined in Section 2.1 (Responder versus Non-responder - 1/0) at the clinical location of implanted RV and LV pacing sites constituted the target variable for classifier training. Supplementary Table S1 details the complete feature set comprising all three index categories, providing a comprehensive representation of the clinical, anatomical, and electrophysiological determinants of CRT response.

#### 2.3.3 Machine Learning Classifier Development

The ML pipeline inside the CDSS had three stages: (1) nested hyperparameter optimization; (2) independent performance evaluation through LOO-CV; and (3) final model training on the full dataset. To avoid data leakage, all preprocessing and feature-selection steps were applied exclusively on the training portion of each fold.

The preprocessing pipeline was applied inside each fold, following these steps. First, features with pairwise Pearson correlation |*r*| ≥ 0.80 were removed (see Figure S2 in the Supplementary). Ordered categorical variables (e.g., NYHA class) were encoded using ordinal encoding. To handle class imbalance (40% responders, 60% non-responders), SMOTENC oversampling was applied [34, 35]. For feature selection, Recursive Feature Elimination (RFE) or Univariate Selection (SelectKBest) was used, with the number of retained features (*k*=2–20) treated as a hyperparameter. Four scaling options were evaluated: StandardScaler, MinMaxScaler, Yeo–Johnson, and Quantile Transformer [36]. The Quantile Transformer consistently performed best, because its robustness to non-Gaussian distributions in our dataset (*n*=74).

For model selection, nested cross-validation was used, with LOO-CV in the outer loop and 5-fold stratified CV in the inner loop. Bayesian optimization via the Tree-structured Parzen Estimator (Optuna, 100 trials) [37] jointly tuned: SMOTENC application, feature-selection method and *k*, scaler type, and classifier type. The F1-score served as the optimization objective. Five classifiers were compared: Support Vector Machine (SVM), Random Forest (RF), Logistic Regression (LR), Linear Discriminant Analysis (LDA), and Gradient Boosting Classifier (GBC). For SVM, *C* ∈ [10*^−^*^3^, 10^2^], kernel type (linear, RBF, polynomial), and kernel parameters were optimized. The best configuration used SMOTENC, SelectKBest with 11 features, Quantile Transformer, and SVM with RBF kernel.

LOO-CV was used as the primary performance estimate given the limited cohort size (*n*=74). The classifier was calibrated using Platt scaling with 3-fold CV. The final model was trained on the full dataset using the optimal configuration. Pairwise correlated features (|*r*| ≥ 0.8) were removed prior to training (see Supplementary Figure S2). The classification threshold was determined by applying Youden’s J statistic to the out-of-fold predicted probabilities from LOO-CV, yielding an optimal threshold of 0.5.

Model stability was estimated via bootstrap resampling (*n*=200), providing 95% CI for AUC while holding hyperparameters constant. Statistical significance was confirmed by permutation testing (*n*=200), where response labels were shuffled and LOO-CV AUC was recomputed. The proportion of permutations achieving AUC ≥ the observed value provided an empirical *p*-value.

#### 2.3.4 Coronary Sinus Anatomy Processing and Candidate Site Selection

To constrain LVPS optimization to clinically implantable locations, a semi-automated pipeline was developed to define candidate pacing sites within each patient’s CS anatomy. First, a 3D centerline was extracted from the smoothed triangular surface mesh of the CS lumen via skeletonization (Figure 2, 1). A custom Python algorithm marked each venous branch of the CS tree. Then, a clinical expert did manual filtering to exclude the posterior interventricular vein and the main CS body, keeping only tributaries that met the implantation rules: diameter ⩾ 2 mm and ostial angle ⩾ 30 degrees (Figure 2, 2). Tributaries with ostial diameter *<* 2 mm (below the body diameter of standard LV leads) and takeoff angle *<* 30 degrees were excluded based on institutional implantation experience as inaccessible for conventional lead delivery. Accepted centerline segments were approximated with splines, and candidate LVPS were sampled at 4–5 mm intervals, yielding a discrete set of clinically constrained testing sites (Figure 2, 3). For LVPS optimization, electrophysiological simulations were run at all candidate sites, LVPS-related features were extracted, and the ML classifier was applied to each LVPS dataset to compute a response likelihood score (ML-score). All candidate sites were labelled on the 3D CS model according to their ML-score; sites with ML-score ⩾ 0.5 were marked as predicting positive response to BiVP (binary outcome is 1), and the site yielding the maximum ML-score was designated as the in-silico optimal LVPS (LVPS*_opt_*). If the maximum ML-score across all CS sites was *<* 0.5, the patient was classified as unlikely to respond to conventional BiVP (binary outcome is 0) regardless of lead placement within the accessible CS anatomy.

**Fig. 2.**
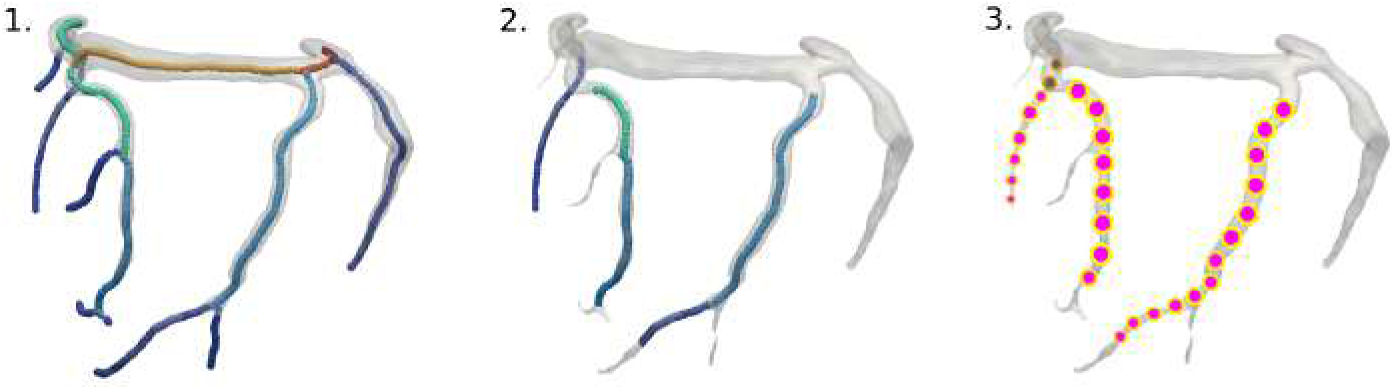
Identification of clinically implantable LVPS candidates within the CS venous tree. Three-stage pipeline: (1) Anatomical centerline extraction from 3D CS geometry reconstruction, with segments labelled by parent vein. (2) Medical expert selects anatomically accessible tributaries, excluding the main CS body and sites with unfavourable angulation or diameter. (3) Site sampling across spline approximation of selected segments with candidate LVPS sampled at 4–5 mm intervals.

**Fig. 3.**
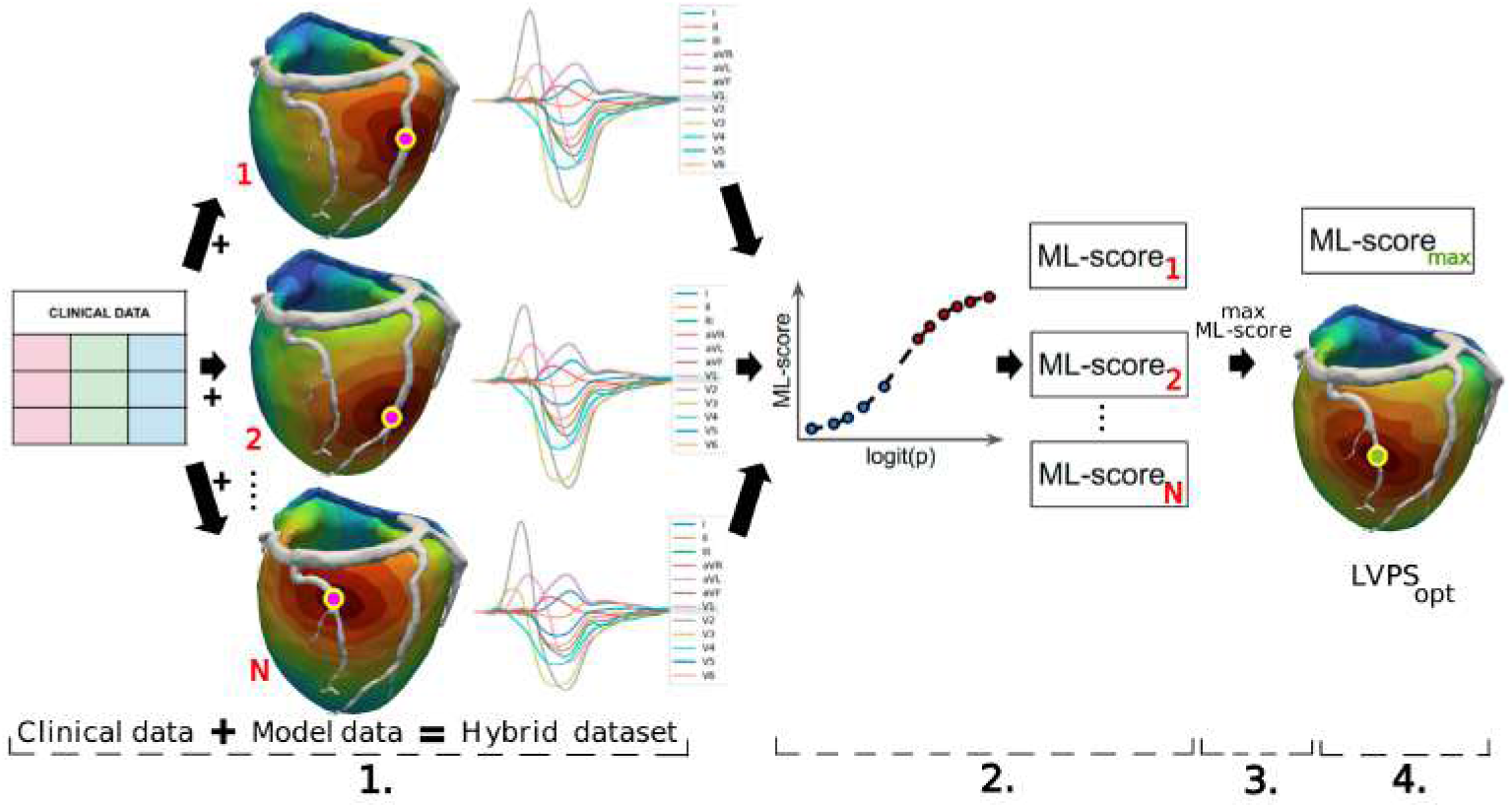
ML-guided LVPS optimisation algorithm within the CDSS framework. Four sequential stages: (1) patient-specific feature extraction from clinical data and electrophysiological simulations at candidate CS sites; (2) ML-score computation for each site using the pre-trained ML classifier; (3) spatial labelling of candidate sites by predicted response probability; (4) identification of the optimal implantable site (LVPSopt) as the highest-scoring accessible location.

#### 2.3.5 Model Explainability: SHAP Analysis

To provide a clinically interpretable rationale for each CDSS recommendation, a SHAP (SHapley Additive Explanations) analysis was implemented as an integral component of the framework’s output layer. SHAP is not a post-hoc validation step. Rather, it is the mechanism through which the system translates the basis of its predictions to the clinician. For each patient and candidate LVPS, SHAP values were computed for the trained ML classifier using the kernel SHAP estimator. A positive SHAP value for a feature indicates that its value at the given LVPS increases the predicted probability of response, while a negative value indicates a reduction.

Two levels of SHAP output were generated. At the cohort level, a SHAP summary plot (Figure 4A) ranks features by mean absolute SHAP value across all patients and candidate LVPSs, identifying globally important predictors. At the patient level, SHAP waterfall plots were generated for five reclassified non-responders. These plots show the decomposition of the shift from a negative to a positive ML score at the model-suggested LVPS relative to the clinical LVPS, feature by feature.

**Fig. 4.**
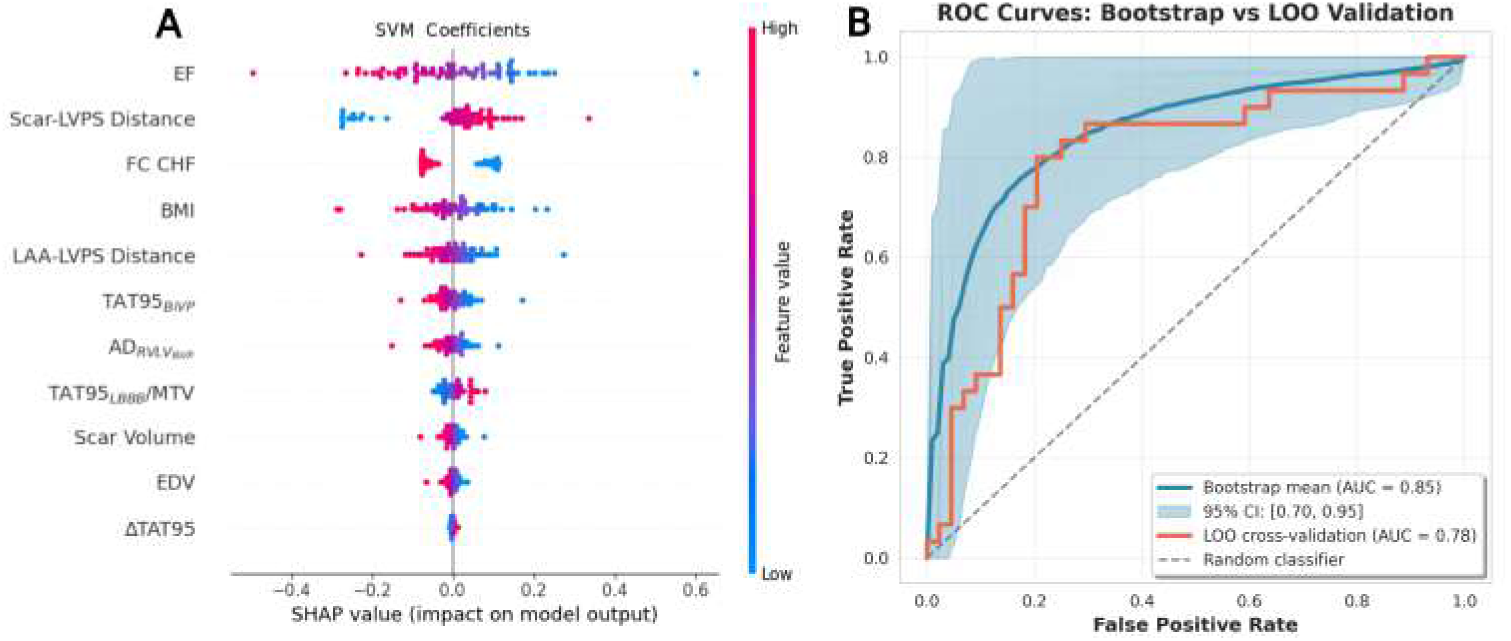
Performance interpretation of the best ML classifier (SVM) for CRT response prediction trained on the entire dataset from 74 patients. (A): SHAP Summary Plot displaying the selected features ranked in descending order of importance for predicting CRT response probability (ML-score). SHAP values on the x-axis indicate the direction of feature influence (positive or negative impact) on the classifier output, while the colormap represents the feature value effect on the ML-score. (B): Comprehensive ROC analysis comparing bootstrap resampling and LOO-CV. ROC curves from 200 bootstrap iterations (light blue) with mean curve (dark blue) and 95% confidence interval. The bootstrap analysis demonstrates model performance with a mean AUC=0.85, 95% CI [0.70-0.95]. The orange line shows LOO-CV performance with an AUC=0.78.

### 2.4 Software

Cardiac electrophysiology was simulated using a custom software implemented in Python using the FEniCS finite element library [38]. Anatomical processing included centerline extraction from the coronary sinus geometry performed with PyVista [39], and assignment of myocardial fiber orientations, UVC, and AHA segment labels using the third-party software [27]. Fibrosis regions were integrated into the volumetric heart mesh through a custom algorithm utilizing GMSH [25], following our previously established methodology [17, 40]. The ML pipeline for CRT outcome classification, feature selection, cross-validation, and ROC-AUC analysis was implemented using scikit-learn [41].

### 2.5 Statistical Analysis

Statistical analysis was performed using Python with the SciPy library [42]. Categorical data are presented as frequencies and percentages. Continuous data are presented as mean ± standard deviation. The Shapiro-Wilk test was used to assess the normality of the distribution. Given the sample size and frequent non-normality of biomedical data, non-parametric tests were primarily employed for group comparisons. The Mann-Whitney U test was used for comparisons between two independent groups (e.g., responders *vs* non-responders) for continuous variables. Wilcoxon signed-rank test was used for paired comparisons within the same group for quantitative data, and McNemar’s test for qualitative data. Pearson’s chi-square test was used for categorical variables. A two-sided p-value of less than 0.05 was considered statistically significant. Statistical evaluation of ML classifier performance, including bootstrap confidence intervals and permutation testing, is described in Section 2.3.3.

## 3 Results

### 3.1 Cohort Characteristics

The population cohort contained 74 patients who underwent CRT implantation with a mean age of 63.6±6.6 years, and a male predominance (70.3%). In the entire cohort, 30 (40.5%) patients were defined as responders according to the CRT response criteria (ΔLVEF*>*10%). The remaining 44 (59.5%) non-responders demonstrated significantly higher left ventricular ejection fraction (LVEF) values (27.8±5.9% *vs* 22.5±4.8%, p*<*0.01) and lower end-systolic volume (ESV) (197.1±71.6 ml *vs* 224.4±59.8 ml, p*<*0.05) at baseline LBBB activation. CT/MRI-derived indices showed that responders had greater Scar-LVPS Distance (40.36±25.5 mm *vs* 25.5±23.9 mm, p*<*0.05) and smaller fibrosis volumes (34.7±30.2 ml *vs* 47.2±7.6 ml, p*<*0.05) as compared with non-responders. Univariate logistic regression analysis (see Table S2 in the Supplementary) confirmed these associations and additionally identified the distance from the LVPS to the late activation area (LAA-LVPS Distance) as a significant predictor of CRT response (odds ratio 0.98 [0.96-1.00], p=0.040). Other model-derived indexes did not show statistically significant differences between responders and non-responders (see Supplementary Table S1). However, no single feature provided adequate discriminative power on its own, which motivated us to integrate a multivariable ML approach.

#### 3.1.1 CS patient group characteristics

In the CS group, six out of 19 (31.6%) patients were classified as CRT responders according to the ΔLVEF*>*10% response criteria, with an average ΔLVEF of 18±8% and LV ESV reduction (ΔESV) of -39±12% (see Table 1). The majority of the CS group (13 patients, 68.4%) was classified as CRT non-responders, with an average ΔLVEF of 6.7±1.6% (p*<*0.05) and a ΔESV of -19±2% (p*<*0.001).

For the CS group, we created a detailed 3D heart model incorporating myocardial fibrosis obtained through MRI-based segmentation, as well as CS tributaries obtained through CT scans. For each patient in the CS group, the right and left ventricular pacing sites (RVPS and LVPS) were manually defined using CT scans. For most patients, the RVPS was located in the apical area of the RV. The LVPS was located on the anterior, posterior, or lateral walls, with additional sectioning into the basal, midventricular, and apical parts. We used the AHA 17-segment LV model for a more detailed description of the LVPS position (Table 3). In patients from the CS group, the LV lead was positioned through the CS vein on the lateral LV wall in 78.9% of patients and the anterior wall in 21.1% of patients. The LVPS was midventricular in 63.2% of patients and basal in 31.6% of patients. As for the non-CS group, the frequency of LVPS placement into the lateral wall was not significant between the responders and non-responders in the CS group.

The average myocardial scar burden in the CS subgroup was 24±15% of LV volume. Implantation of the LVPS into scar tissue was observed in 1 responder (16.7%) and 5 non-responders (38.5%); this difference was not statistically significant (p=0.605), however, this subgroup comparison was substantially underpowered (6 vs 13 patients). While the distance from the LV lead to the nearest scar (Scar-LVPS Distance) was numerically greater in responders, this trend did not reach statistical significance within this specific subgroup (p=0.196).

Simulations of biventricular pacing in personalized models provided electrophysiological parameters, such as total activation time (TAT95), change in total activation time (ΔTAT95), and interventricular delay, among others (see Table S1 in the Supplementary). There was no statistically significant difference between responders and non-responders within the CS group for any of these simulated indices (p*>*0.05); however, these comparisons were limited by the small subgroup size (n=6 vs n=13).

#### 3.1.2 Comparative Analysis of CS group versus Non-CS group

Patients in the CS group had a significantly smaller QRSd at baseline ventricular activation (182.0±18.6 ms *vs* 191.7±23.7 ms, p*<*0.05), end-diastolic volume (EDV, 250.7±77.6 ml *vs* 291.9±82.2 ml, p*<*0.05), end-diastolic diameter (EDD, 69.5±7.0 mm, *vs* 73.8±7.4 mm, p*<*0.05), and substantially shorter Scar-LVPS Distance (18.7±24.8 mm *vs* 35.9±24.4 mm, p*<*0.01). Notably, there was a substantial divergence in myocardial tissue volumes between subgroups. The CS group showed markedly lower total myocardial volume (240.1±89.2 ml *vs* 358.8±141.6 ml, p*<*0.01) and left ventricular tissue volume (137.2±66.4 ml *vs* 205.3±98.0 ml, p*<*0.01). Dyssynchrony indices (*AD_RV LV_*) exhibited distinct patterns. The CS group had greater baseline interventricular dyssynchrony LBBB (109.2±33.9 ms *vs* 83.6±34.9 ms, p*<*0.01) but experienced more pronounced relative reduction during biventricular pacing (−0.91±0.22% *vs* -1.12±0.39%, p*<*0.05).

These systematic anatomical and electrophysiological differences between the Non-CS and CS cohorts indicated a distributional shift in the feature space. To mitigate this, all features were transformed using a Quantile Transformer within each cross-validation fold, which maps heterogeneous marginal distributions to a common reference distribution. While this preprocessing step does not constitute formal domain adaptation, it reduces the influence of scale and distributional differences between subgroups on classifier performance.

### 3.2 Performance of the ML Classifier

As the core predictive component of the CDSS, we trained an ML classifier for the prediction of CRT-response defined as ΔLVEF*>*10% on the hybrid dataset. We considered several ML models: LR, SVM, RF, LDA, and GBC. As a result, the SVM showed a maximal F1-score during LOO-CV and nested hyperparameter optimization described in our ML pipeline 2.3.3.

The trained SVM classifier substantially outperformed the reference Feeny et al. clinical calculator [24] on the same cohort using identical response definition: accuracy 0.78 vs 0.58, F1-score 0.75 vs 0.43.

Under LOO-CV, the classifier achieved AUC = 0.78, accuracy = 0.78, sensitivity = 0.80, specificity = 0.77 (Table 2). Bootstrap analysis (n = 200) yielded mean F1-score of 0.80 (95% CI: [0.64–0.91]) and AUC = 0.85 (95% CI:[0.70–0.95]), confirming stability across data perturbations (Figure 4B). Permutation testing confirmed performance above chance (p *<* 0.05) for ROC AUC. The non-significant p-value for the F1-score (p = 0.260) suggests that while the overall discriminative ability is stable, the precise classification threshold may be sensitive to data perturbations within this sample size. This represents a limitation that should be addressed with a larger validation cohort. Table 2 shows characteristics of the SVM classifier performance.

**Table 2.**
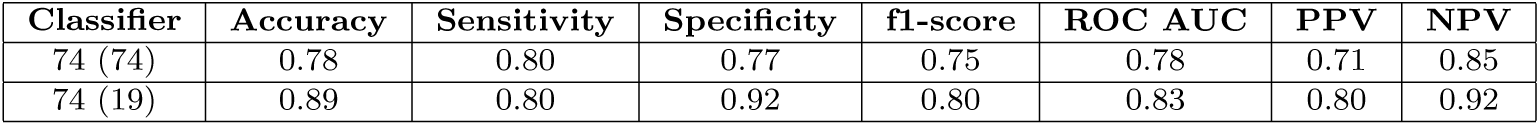
Performance metrics of the SVM classifier obtained through leave-one-out (LOO) cross-validation. Top row: metrics computed across all 74 patients in the cohort. Bottom row: metrics computed specifically on the subgroup of 19 patients with extensive fibrosis and coronary sinus anatomy (CS group), using only their LOO predictions.

The 11 selected features are shown in Figure 4A, ranked by mean absolute SHAP value. Three of the 11 most important selected features are baseline clinical features at LBBB activation pattern before CRT implantation. Baseline LVEF is the most significant feature, having the greatest negative impact on the ML-classifier score (the higher the baseline LVEF, the lower the score predicting LVEF improvement). The next two features are the AHA functional class (FC) of CHF and body mass index (BMI), both of which have a negative impact on the ML score.

The three most important model-derived features were the Scar-LVPS distance, LAA-LVPS distance, and TAT95 under BiVP. Note that the latter feature depends on the LVPS position, which can affect the ML score. Scar-LVPS positively influences the ML score. The distance from the LVPS to the baseline late activation area (LAA) is the second most important model-derived index in the ML model.

LAA-LVPS distance ranked as the second most important model-derived index. The most important remaining indices were the model-driven electrical dyssynchrony indices computed at baseline LBBB (TAT95*_LBBB_*/MTV) and during BiVP (TAT95*_BiV P_*, AD*_RV LVBiV P_*), MRI-derived scar volume, echocardiography-derived end-diastolic volume (EDV) of LV measured at baseline LBBB, and the relative changes of TAT95 (ΔTAT95).

### 3.3 Core finding: CDSS output for the CS pilot cohort

#### 3.3.1 Patient stratification and LVPS optimization

We applied our developed CDSS framework to the 19-patient CS pilot cohort for providing two clinically relevant stratification outputs.

Individual ML-score values and AHA segment classifications for all 19 patients at both clinical and optimal LVPS are provided in Table 3. Among the 6 clinical responders, the ML-score did not increase meaningfully at any alternative CS site (maximum increment 0.05). The two false-negative cases (Patients 05 and 22) remained unimproved. The framework did not identify alternative sites that would correct these false-negative predictions, suggesting that the misclassification is driven by patient-level features rather than LVPS location.

**Table 3.** Classification of patients from the CS group for the clinical and optimal LVPS within the CS tributaries. Shown are: Responder/Non-responder coded as 1/0, number of AHA LV segments according to the 17-segment AHA model, ML-score value depending on the LVPS location, Positive/Negative CRT response prediction coded as 1/0 depending on the ML-score above/below the threshold of 0.5, Concordant CS tributary for the clinical and optimized LVPS coded as 1/0. The bold rows highlight patients classified as true negatives at the clinical LVPS, who were re-classified as positive at the optimal LVPS.

|  | Observed<br>clinical<br>outcome | Clinical LVPS |  |  | Optimal LVPS |  |  |  |
| --- | --- | --- | --- | --- | --- | --- | --- | --- |
| Patient<br>ID | Responder/<br>Non-responder | AHA LV<br>segment | ML-score | Positive/<br>Negative | Concordant<br>CS tributary | AHA LV<br>segment | ML-score | Positive/<br>Negative |
| 01 | 1 | 12 | 0.75 | 1 | 1 | 6 | 0.76 | 1 |
| 02 | 1 | 6 | 0.98 | 1 | 1 | 6 | 0.98 | 1 |
| 05 | 1 | 12 | 0.06 | 0 | 1 | 5 | 0.08 | 0 |
| 09 | 1 | 6 | 0.73 | 1 | 1 | 7 | 0.78 | 1 |
| 22 | 1 | 12 | 0.33 | 0 | 0 | 1 | 0.37 | 0 |
| 33 | 1 | 16 | 0.55 | 1 | 1 | 16 | 0.56 | 1 |
| **03** | **0** | **7** | **0.30** | **0** | **0** | **5** | **0.76** | **1** |
| **04** | **0** | **6** | **0.30** | **0** | **1** | **1** | **0.52** | **1** |
| 07 | 0 | 6 | 0.33 | 0 | 1 | 12 | 0.40 | 0 |
| 10 | 0 | 6 | 0.30 | 0 | 1 | 6 | 0.30 | 0 |
| **14** | **0** | **12** | **0.38** | **0** | **0** | **1** | **0.57** | **1** |
| 15 | 0 | 5 | 0.26 | 0 | 1 | 12 | 0.43 | 0 |
| 16 | 0 | 11 | 0.77 | 1 | 0 | 1 | 0.86 | 1 |
| 17 | 0 | 12 | 0.13 | 0 | 1 | 6 | 0.37 | 0 |
| **20** | **0** | **11** | **0.26** | **0** | **1** | **6** | **0.59** | **1** |
| **21** | **0** | **12** | **0.45** | **0** | **0** | **16** | **0.56** | **1** |
| 23 | 0 | 12 | 0.30 | 0 | 0 | 6 | 0.34 | 0 |
| 24 | 0 | 7 | 0.32 | 0 | 1 | 1 | 0.38 | 0 |
| 30 | 0 | 11 | 0.14 | 0 | 0 | 1 | 0.32 | 0 |

For 8 of 13 clinical non-responders the CDSS framework predicted no accessible CS site with ML-score ≥ 0.5. It indicates that conventional BiVP is unlikely to produce an expected response regardless of lead placement within the available CS anatomy (Table 3). These patients were classified as unlikely to respond to conventional BiVP at any accessible CS site.

For the remaining 5 clinical non-responders (Patients 03, 04, 14, 20, 21) the framework predicted a positive response at an alternative accessible LVPS within the CS with ML-score ≥ 0.5. For brevity, these cases are referred to as ”reclassified to virtually positive”, although this reflects an in silico prediction, not a clinically validated outcome. In these patients, the ML-score increment from clinical to optimal LVPS ranged from 0.11 to 0.46 (Table 3). In all five clinical non-responders, who were reclassified as virtually positive, the optimized LVPS were located in alternative AHA LV-segments as compared with clinical LVPS (Table 3). Of note, in a majority of the patients, the optimized LVPS was located in the basal segments (4 of 5), and lateral segments were more frequent than other (3 of 5).

Across the entire CS cohort, we found a higher median ML-score in responders vs non-responders for baseline LBBB in the CS group (0.64 [0.26-0.80] *vs* 0.30 [0.25-0.35], p=0.044) as expected. The mean ML-score at the optimal LVPS was significantly higher than at the clinical LVPS (0.52 ± 0.23 *vs* 0.40 ± 0.25, p *<* 0.001). In 12 of 19 patients (63%), the optimal LVPS was located in the same CS tributary as the clinical LVPS, though in a different AHA LV segment in 16 of 19 patients. Optimized LVPS were significantly more remote from fibrosis regions than clinical LVPS (Scar-LVPS distance: 19 [6–37] vs 3 [0–35] mm, p = 0.048). No significant difference in LAA-LVPS distance was observed between optimal and clinical LVPS (45 [30–63] vs 47 [38–63] mm, p = 0.778).

#### 3.3.2 Illustrative case studies: visualization of CDSS outputs

Three representative examples shown in Figure 5 visualize CDSS outputs across different clinical outcomes. The three examples were selected to demonstrate potential distinct CDSS outputs, not to illustate clinical efficacy at model-suggested sites. Each sphere on the CS tributaries indicates an ML-score value predicted at LVPS located at this CS point, and shades of red denote positive predictions (ML-score ≥ 0.5), while blue shades denote negative predictions (ML-score *<* 0.5).

**Fig. 5.**
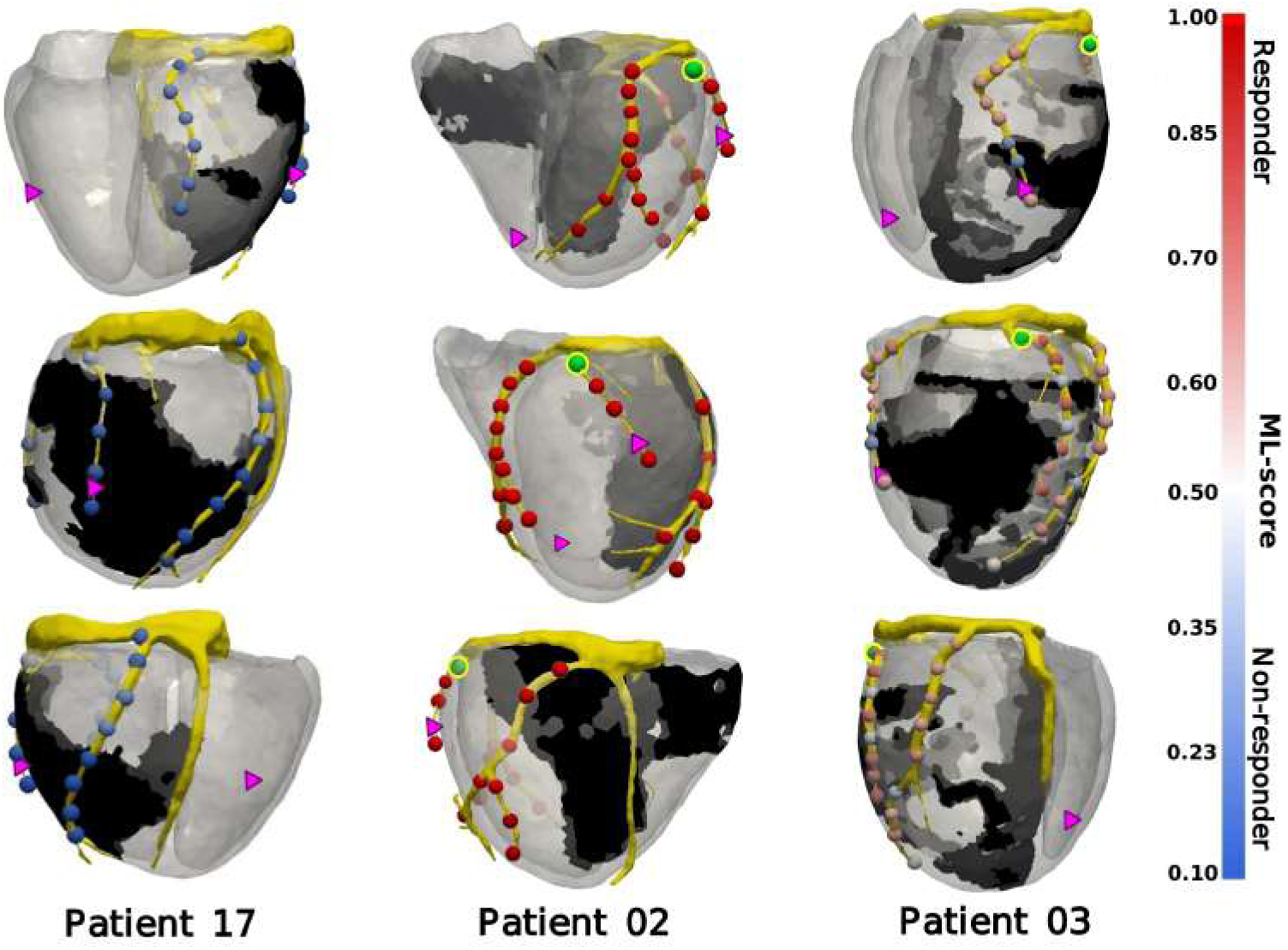
CDSS outputs and spatial mapping of ML-derived CRT response probability. Colored spheres represent potential left ventricular pacing sites (LVPS) within the coronary sinus (CS) anatomy, with colors indicating the ML-score (probability of response). Red spheres denote positive predictions (ML-score *≥* 0.5), while blue spheres denote negative predictions (ML-score *<* 0.5). Pink triangles indicate the actual clinical LVPS and RVPS positions derived from CT imaging. Green spheres highlight the optimal LVPS (site with the maximum ML-score). Black regions represent myocardial fibrosis, and the yellow structure depicts the CS geometry. Examples (from left to right) include: (Patient 17) a clinical non-responder; (Patient 02) a clinical responder; and (Patient 03) a clinical non-responder predicted by the SVM classifier to achieve response if re-positioned to the identified optimal CS site.

Left panel shows a negative case with no accessible CS LVPS predicting positive CRT outcome (Patient 17, clinical non-responder, with an 8% LVEF improvement). All candidate LVPS within the CS tributaries are classified as negative with ML-scores *<* 0.5 (clinical 0.13; maximum 0.37). The framework correctly classified this patient as unlikely to benefit from conventional BiVP, regardless of LVPS placement. This outcome demonstrates the CDSS’s stratification function of redirecting clinical decisions towards alternative strategies rather than lead repositioning.

Central panel demonstrates a positive case with all virtually positive CS LVPS. Clinical LVPS is already located near the optimal position predicted by the ML-model. All candidate LVPS received ML-score ≥ 0.5. This patient 02 is a clinical responder, with a 14% LVEF improvement. The maximum ML-score of 0.98 matched the clinical site’s score. The optimal LVPS was located in the same vein as the clinical site, but in an alternative basal LV segment. This illustrates the CDSS confirmation function, validating an existing clinical decision and providing spatial guidance for fine-tuning within the same tributary.

Right panel illustrates a potentially optimized case where an alternative virtually positive LVPS was identified in a different CS tributary (Patient 03, non-responder, with a 5% LVEF improvement). The clinical LVPS (AHA segment 7, ML-score 0.30) was implanted into the fibrosis boundary (Scar-LVPS distance 0 mm). The CDSS identified an alternative site in a different tributary (AHA segment 5, ML-score 0.76), which was located 20 mm apart from the fibrosis boundary. The SHAP explanation attributed the reclassification primarily to increased Scar-LVPS distance and a reduced TAT95 under BiVP. This mechanistic rationale can be communicated to the implanting electrophysiologist in standard electrophysiology terms.

### 3.4 Explanation of CRT outcome prdictions based on SHAP analysis

The developed framework uses SHAP analysis at both the population and patient levels. For each of the five clinical non-responders reclassified as virtually positive, the individual SHAP waterfall plots (Figure 6) provide a feature-by-feature decomposition of the shift from negative to positive CRT-outcome prediction. The SHAP explanations utilized the 11 most important features selected by the ML classifier for examination of their contribution to the individual CRT prediction.

**Fig. 6.**
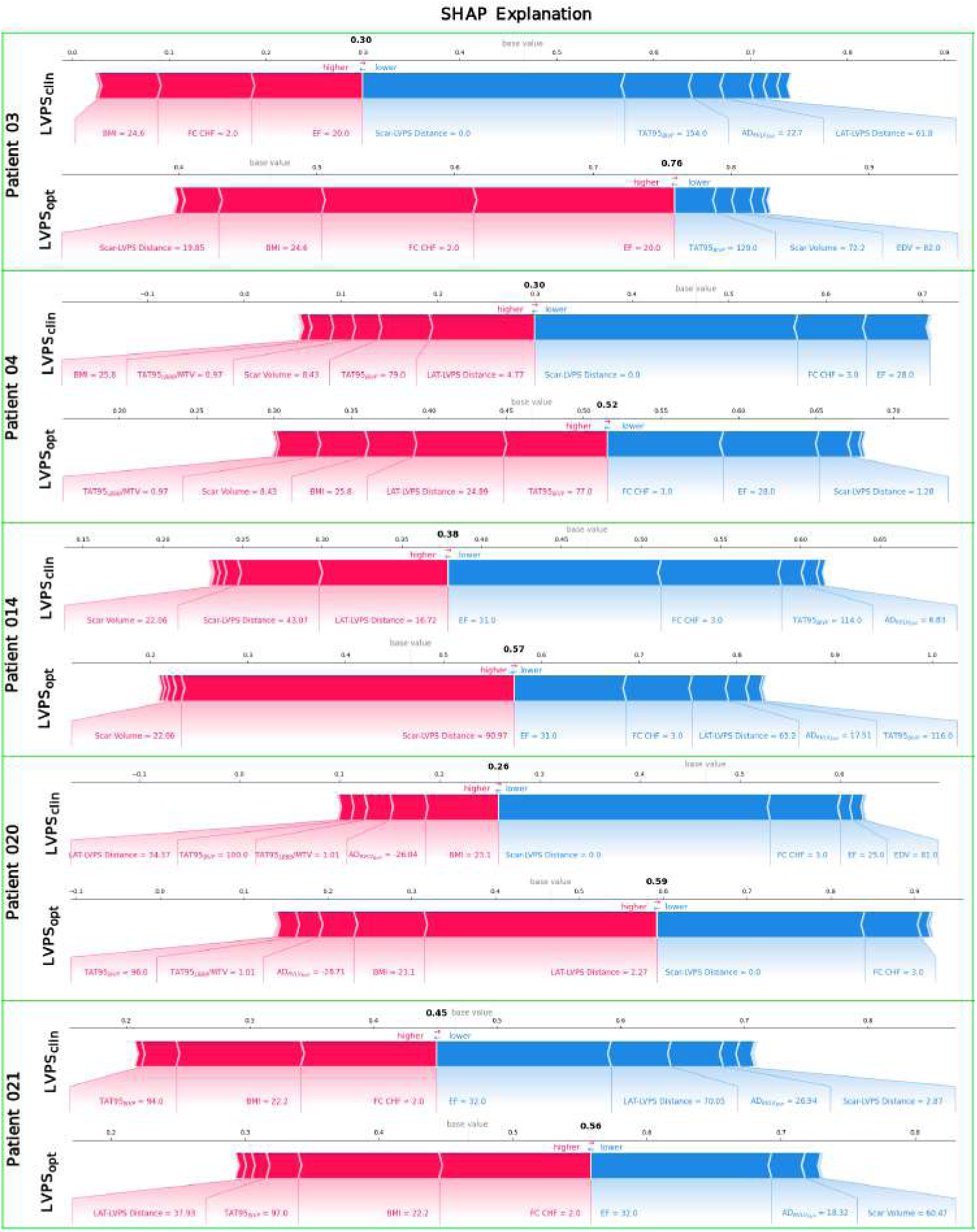
An individual SHAP explanation of the input variable contributions to the ML score generated by the best SVM classifier of CRT response inside the CDSS. Shown is an analysis for each of five clinical non-responders classified as true negative on the input data for the clinical LVPS (upper lines, ML-score *<* 0.5), and re-classified into positive on the input data for the optimal LVPS (lower lines, highest ML-score *≥* 0.5). The diagrams indicate input variables of a positive impact on the ML score (rose arrows; the longer the arrow the higher the impact), and variables of a negative impact on the score (blue arrows). The explanation clearly illustrates a mechanistic, data-driven multivariable effect of LVPS location on the CRT response prediction.

We found that the distance from the optimal LVPS to the scar area (Scar-LVPS distance) increased in the CS group compared to the clinical LVPS. For the five reclassified clinical non-responders, a notable increase in Scar-LVPS distance was predicted for two patients (Patients 03 and 14), while no significant change was observed for the other three patients (Figure 6). For the same 5 patients, the distance from the optimal LVPS to the late activation area (LAA-LVPS distance) decreased for Patient 03, Patient 20, Patient 21, while in contrast it increased in patients Patient 04, Patient 14 (Figure 6). Both these distances (Scar-LVPS and LAA-LVPS) were shown as the most important model-derived features contributing to the ML-score value (Figure 4). However, no correlation was observed between changes in each of these features and the increase in ML-score due to the LVPS optimization (ΔML-score *vs* ΔScar-LVPS Distance; r=0.139, p=0.571; ΔML-score *vs* ΔLAA-LVPS Distance; r=0.121, p=0.622). We found no significant reduction in total activation time (TAT95 BiVP) of the ventricles under BiVP at the optimized LVPS versus the clinical LVPS in these 5 patients. The activation delay of the LV *vs* RV (AD*_RV LV_*) at BiVP with the optimized LVPS varied from patient to patient and, in some cases, changed sign compared with the clinical LVPS (Table 4).

**Table 4.** Comparison of eight most important input clinical and model-derived variables fed to the SVM classifier for CRT response prediction in five non-responders, who were classified as true negative (ML score *<* 0.5) at the clinical LVPS and reclassified as positive (highest ML score *≥* 0.5) at the optimal LVPS.

| Patient ID | LVPS | BMI, kg/m<sup>2</sup> | EF, % | FC CHF, (I,II,III) | Scar-LVPS Distance, mm | LAA-LVPS Distance, mm | TAT95 LBBB, ms | TAT95 BiVP, ms | AD<sub>RV LV</sub> BiVP, ms |
| --- | --- | --- | --- | --- | --- | --- | --- | --- | --- |
| 03 | clinical | 24.6 | 20 | 2 | 0 | 62 | 211 | 154 | 23 |
| 03 | optimal | – | – | – | 20 | 48 | – | 129 | -7 |
| 04 | clinical | 25.8 | 28 | 3 | 0 | 5 | 156 | 79 | -10 |
| 04 | optimal | – | – | – | 1 | 25 | – | 77 | 4 |
| 14 | clinical | 28.0 | 31 | 3 | 43 | 17 | 145 | 114 | 7 |
| 14 | optimal | – | – | – | 91 | 65 | – | 116 | 18 |
| 20 | clinical | 23.1 | 25 | 3 | 0 | 34 | 214 | 100 | -26 |
| 20 | optimal | – | – | – | 0 | 2 | – | 96 | -29 |
| 21 | clinical | 22.2 | 32 | 2 | 3 | 70 | 163 | 94 | 27 |
| 21 | optimal | – | – | – | 7 | 38 | – | 97 | 18 |

The SHAP analysis revealed that the factors most influential in shifting from a negative to a positive prediction varied among the five reclassified clinical non-responders (Figure 6). For Patients 03, 04, and 20, the clinical LVPS was co-localized with the fibrosis area. In these cases, we defined the distance to the fibrosis as 0 and a low ML-score value associated with a negative prediction of the CRT response. Across the five cases, three distinct mechanisms of reclassification were observed: increased distance from fibrosis (Patients 03, 14), decreased distance to the LAA (Patients 04, 20, 21), and reduced dyssynchrony (Patient 21), often in combination. However, in Patient 03, the highest ML-score at the optimized LVPS was improved by a combination of an increased distance to the fibrosis and a reduced TAT95 in BiVP compared to the clinical ML-score. In Patient 04, a positive ML-score was achieved through a combination of a reduced distance to the LAA and a non-zero distance to the fibrosis area. In Patient 20, the optimized LVPS remained within the fibrosis area (zero distance), but a significantly shorter distance to the LAA contributed to the positive prediction. Patient 14 achieved the highest ML score through a substantial increase in distance from the fibrosis area despite a negative increase in distance to the LAA compared to the clinical LVPS. Finally, Patient 21 switched from a negative to a positive CRT response prediction due to a decrease in distance from the optimized LVPS to the LAA and a shorter intraventricular activation delay (AD*_RV LV_*).

## 4 Discussion

### 4.1 Patient-specific guidance over multi-variable heuristics

This study demonstrates that personalized modelling using digital twins and explainable ML can be utilized in clinical decision support tools evaluating CRT outcomes. We developed a new digital framework that enables 3D visualization of accessible CS tributaries merged with ventricular anatomy and geometry of the myocardial fibrosis; evaluation of the ventricular activation and probability of CRT success at various LVPS locations within the CS anatomy; visualization of the CS map indicating LVPS regions of positive and negative response probability, and stratification of CRT candidates with recommendations for optimal LVPS location for potentially positive patients. SHAP analysis provides explanations for ML classifier decision at each LVPS.

The central finding of this study is that selection of the LVPS within the CS is a multivariable problem. It cannot be solved by optimizing any single anatomical or electrophysiological parameter, but requires their joint consideration in the context of patient’s individual decision. Training the classifier on a hybrid dataset containing multi-modal data from patient anatomy, myocardial substrate, and simulated electrophysiology produces guidance that is clinically feasible and can hardly be replaced by any single-variable heuristic. We found no single feature change to be predictive of the change in ML-classifier score induced by the change in the LVPS location. Particularly, we found no significant univariable correlations of the following features with the ΔML-score in the CS group: ΔScar-LVPS distance (r = 0.139, p = 0.571), ΔLAA-LVPS distance (r = 0.121, p = 0.622), ΔTAT95 (r=-0.21, p=0.717), and ΔAD*_RV LV_* (r=-0.18, p=0.104).

As shown in the Table 4, the LVPS-dependent indexes (Scar-LVPS distance, LAA-LVPS distance, TAT95 BiVP, AD*_RV LV_* BiVP) changed to a different extent and even in the opposite directions at the optimal LVPS compared with the clinical LVPS among the five reclassified non-responders. No association was found in either the responders or non-responders subgroups for ΔML-score and other indexes. This finding reflects the multivariable nature of LVPS-dependent CRT response.

Using our multivariable approach, we were able to predict alternative locations of LVPS within available CS tributaries where the ML classifier predicted a positive response to BiVP in five (38%) clinical non-responders in the CS group classified as true-negative at the clinical LVPS position. Current optimization protocols focusing on either the reduction in LAA-LVPS distance [9, 10], or avoiding fibrosis [3, 6], or maximizing QLV delay [7, 8] optimize a single variable from a limited subset of relevant variables without constraining optimization to the patient-specific CS anatomy. Notably, in our patient cohort Scar-LVPS distance ranked higher than LAA-LVPS distance by SHAP importance. This contrasts with the prevailing clinical strategy of targeting the latest-activated area and suggests that avoidance of scar may be a more robust determinant of LVPS-dependent response than proximity to the LAA. Our findings suggest that candidates for CRT may benefit from an optimized LVPS placement within the CS taking onto account a combination of significant parameters which could improve their prognosis.

The advantage of our classifier performance over the Feeny clinical calculator (accuracy 0.78 *vs* 0.58, F1-score 0.75 *vs* 0.43) is unlikely to be explained by the ML algorithm selection. Feeny et al. compared six classifiers, including SVM, on a big (n¿1000) dataset of clinical features and found that all except AdaBoost achieved comparable AUCs (0.70–0.72), demonstrating low impact of algorithm choice on prediction from clinical variables. We believe that the observed advantage of our approach is therefore primarily attributable to the inclusion of simulation-derived features that capture patient-specific predictions on LVPS dependent characteristics unavailable from pre-procedural clinical data. Notably, the restrictive CRT response definition (ΔLVEF *>* 10%) used here was specifically selected to enable a direct comparison with the Feeny calculator, which also used this criterion. The use of the same response definition ensures a direct and fair comparison between the two approaches.

### 4.2 When CS optimization is insufficient: epicardial alternatives and scope of BiVP non-response

For the majority of clinical non-responders in this cohort (8 of 13 non-responders), the framework found no accessible CS site predicted to yield a positive response, regardless of lead placement. If the maximal ML score is *<* 0.5 for the CS geometry, an alternative pacing strategy should be considered, especially if the available CS veins are unsuitable. In these instances, our CDSS can predict alternative LVPS locations on the epicardial and endocardial LV surfaces.

As an exploratory post hoc analysis, we computed ML-score for the CRT response prediction at LVPS located on the epicardial surface at the centers of LV segments according to the 17-segment AHA model (see Table S3 in the Supplementary). In a majority (14 of 19) of patients from the CS group, alternative LVPS locations did not change patient classification *vs* clinical LVPS. For five reclassified clinical non-responders (ML-score *<* 0.5 at clinical LVPS), we found LVPS locations on the epicardial surface that improve prediction with the highest ML-score ≥ 0.5 (see Table S3 in the Supplementary).

Patients 03, 04, 20, and 21 were among the five patients reclassified at the optimized LVPS in the CS veins, and they had similar ML-score values. Interestingly, in each of the four patients, the LVPS with the highest ML-scores were located in different AHA segments. In the case of the fifth Patient 24, which was predicted as negative at any LVPS position within the CS veins, we found that LVPS placed in the center of the 6th AHA basal anterolateral segment predicts a positive response to BiVP, despite the ML-score of 0.5 being on the threshold for a positive prediction.

Our findings show that over half of the non-responders in our CS group had a negative prediction for BiVP response, regardless of the location of the LVPS on the LV epicardial surface, including CS veins. These patients may benefit from alternative CRT designs, such as MPP, LBBAP, or LOT-CRT [43, 44]. The general approach of combining personalized computational models with ML-based outcome prediction can be adapted to evaluate these strategies using corresponding modelling frameworks for conduction system pacing.

### 4.3 Strengths and limitations of the study

This study has a retrospective design, and the model-suggested optimal LVPS locations were not prospectively tested in any patient. Consequently, the reclassification of five non-responders as predicted positive at alternative CS locations represents a hypothesis generated by the model, not a demonstrated clinical benefit. Prospective evaluation for CDSS output is required to assess whether model-guided placement translates into improved response rates.

The main advantage of our approach is combining clinical and simulated data from the digital twins with explainable ML for CRT outcome prediction. It gives a solution for patient stratification, virtual LVPS optimization, 3D visualization and procedure planning simultaneously. Our approach is generally scalable and can be implemented with an extended dataset of clinical data and model-extracted features of different modalities, if available and potentially relevant. Despite the promising results of our pilot study, it should be noted that our dataset for developing the ML classifier was small. Further data accumulation and a prospective study with targeted LVPS placements are required to transform this technique into a real-world decision-making tool for doctors.

An additional practical advantage of the solution we developed is its computational performance. The proposed technology can process from segmented imaging data to CDSS output in less than an hour on consumer-grade devices or a modern cloud-computing instance. However, the current limitation is the semi-automatic process for building patient-specific 3D digital twins. Several new solutions for constructing heart geometry models from clinical images using AI and ML techniques have been suggested [27, 45, 46]. One of the crucial aspects of creating a model is integrating data on fibrosis. Here, AI solutions can also help with automatic implementation [47, 48]. Another important aspect is the automatic personalization of the cardiac electrophysiology models and building non-invasive heart activation maps. Although our CDSS automatically personalizes electrical parameters, different approaches to simulating the electrical conduction system could improve the consistency of our predictions, particularly when a complete LBBB cannot be confirmed from clinical data in patients [49–51].

From a clinical standpoint, we did not simulate varying atrioventricular delays and used a zero RV-LV pacing delay due to the absence of corresponding clinical data. These factors may also affect the CRT response [52–56]. Another limitation is the lack of detailed information on mechanical dyssynchrony during ventricular activation and contraction. Studies [19, 20, 22, 57, 58] have shown that combining measured mechanical indices with model-derived ventricular activation characteristics can significantly predict CRT effectiveness and LV reverse remodelling. Integrating electrical and mechanical model-derived indices is a promising approach to enhancing our CDSS framework. Finally, we only tested one criterion for CRT response, requiring more than a 10% improvement in LVEF. The main reason for this was to compare the performance of our ML classifier with the calculator developed by Feeny based only on clinical data [24]. Many articles consider a reduction in LV ESV of more than 15% to be the criterion for LV remodelling and patient improvement after CRT implantation [1].

## 5 Conclusion

We developed a clinical decision support framework combining clinical data and personalized ventricular simulations. This framework uses explainable ML and digital twins for pre-procedural evaluation of CRT candidates. Integration of patient-specific 3D ventricular and CS anatomy, myocardial fibrosis, and personalized electrophysiological simulation allows us to stratify patients for whom the CRT procedure can give a positive response, identify the alternative LV pacing sites within the CS, and interpret results using SHAP explanations for each recommendation. In 5 of the 13 patients classified as clinically non-responders to CRT, our framework identified alternative pacing sites within the CS tributaries and on the LV surface. These sites were associated with a higher predicted probability of treatment response. These findings support the feasibility of personalized digital twin modelling as a pre-procedural tool for CRT planning. However, further prospective validation in larger multi-center cohorts and regulatory evaluation as medical device software is required before clinical deployment.

## Supporting information

Supplementary Data - SHAP for clinical LVPS

Supplementary Data - SHAP for optimal LVPS

## Data Availability

The anonymized clinical and imaging datasets, along with the model-derived electrophysiological indices generated during the current study, are available from the corresponding author upon reasonable request, subject to institutional data protection policies.

## Acknowledgements

This work was supported by the Russian Science Foundation (grant #24-15-00335). The study received no financial support or funding from XSpline S.p.A. The author acknowledges XSpline S.p.A. for providing access to the 3D engine during her former employment.

## Ethics statements

This retrospective study was conducted in accordance with the Declaration of Helsinki and approved by the Almazov National Medical Research Centre Institutional Ethics Committee (protocol code 203, project No. 18-315-00261/18, approved on 19 November 2018). The research was conducted in compliance with all applicable ethical guidelines for biomedical research involving human subjects. All patient data were anonymized and handled in accordance with institutional data protection policies. Written informed consent was obtained from all participants.

## Supplementary data

Supplementary material related to this article can be found in the files Supplementary-materials.pdf, LVPS-clinical.pdf, and LVPS-ptimal.pdf.

## Author contributions

OS and AB conceived and designed the study. SZ, VS, MB, and DL collected the clinical and imaging data; SZ and MB performed the initial preprocessing. SZ, MB, and AB carried out CT/MRI segmentation and anatomical model construction. AB performed full anatomical model post-processing, developed the finite-element models, assigned infarction regions, conducted computational simulations for LBBB and BiV pacing, implemented the machine-learning models, analyzed the data, and interpreted the results, serving as the primary contributor to the computational and analytical components of the study. SK developed the Purkinje system model. TC and SK contributed to clinical data analysis, interpretation of results, and preparation of descriptive statistics. OS provided major conceptual guidance, supervised all stages of the project, and contributed substantially to the analysis of the results and manuscript writing and editing. OS, AB, TC, and SK drafted significant portions of the manuscript. All authors reviewed and approved the final version of the manuscript.

## Abbreviations

CRT: Cardiac resynchronization therapy
CS: Coronary sinus
CDSS: Clinical decision support system
LV: Left ventricle
RV: Right ventricle
LVPS: Left ventricular pacing site
LBBB: Left bundle branch block
BiVP: Biventricular pacing
ML: Machine learning
SHAP: SHapley Additive Explanations
SVM: Support vector machine
LOO-CV: Leave-one-out cross-validation

