## Supplementary Figures and Tables for "Personalized planning of cardiac resynchronization therapy through integration of coronary sinus geometry, clinical data, digital twins, and machine learning: visualization, stratification, and optimization"

### Supplementary materials

March 24, 2026

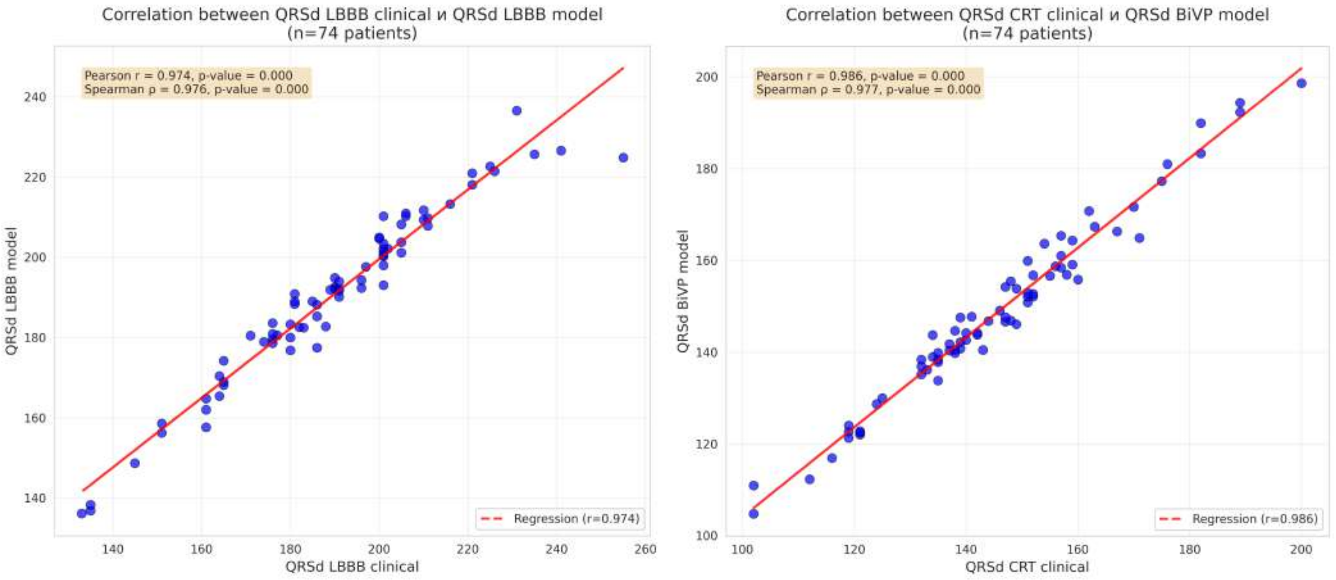


Figure S1: Left panel: Pearson correlation coefficient between QRSd LBBB clinical and QRSd LBBB model. Right panel: Pearson correlation coefficient between QRSd CRT clinical and QRSd BiVP model. The dots represent QRSd values for individual patients.

### 1 Image acquisition and preprocessing

CT imaging was acquired on a Somatom Definition 128 scanner (Siemens Healthcare, Germany) using ECG-gated, contrast-enhanced protocol optimized to visualize ventricle chambers and the CS venous tree. DICOM files were imported into 3D Slicer software, version 5.8.1 (Pieper *et al.*, 2004) to reconstruct 3D torso, CS and ventricular geometries. LGE MRI imaging (INGENIA 1.5 T, Philips) with Late gadolinium enhancement (Gadovist or Magnevist) was performed pre-procedurally to detect fibrosis areas in the myocardium. All DICOMs were converted to NIfTI and underwent bias-field correction (when indicated) and non-local means denoising. Then LGE MRI DICOM files were segmented manually by a clinical expert in Inobitec DICOM Viewer Pro version 2.12 Software (Dubai, UAE) to delineate fibrosis and to match the 3D reconstruction with the anatomy of the ventricles.

### 2 Model-derived indexes

For both the left bundle branch block (LBBB) and biventricular pacing (BiVP) simulations, we calculated ventricular electrical activity using established metrics from our previous work (Khamzin *et al.*, 2021). The total activation time (time for 95% of myocardium to activate) of the ventricles (TAT95), QRS complex duration (QRSd), and three indices of ventricular electrical dyssynchrony were calculated:

$TAT95 = t_{95} - t_0$

$QRSd = max\{T_S - T_Q\}$

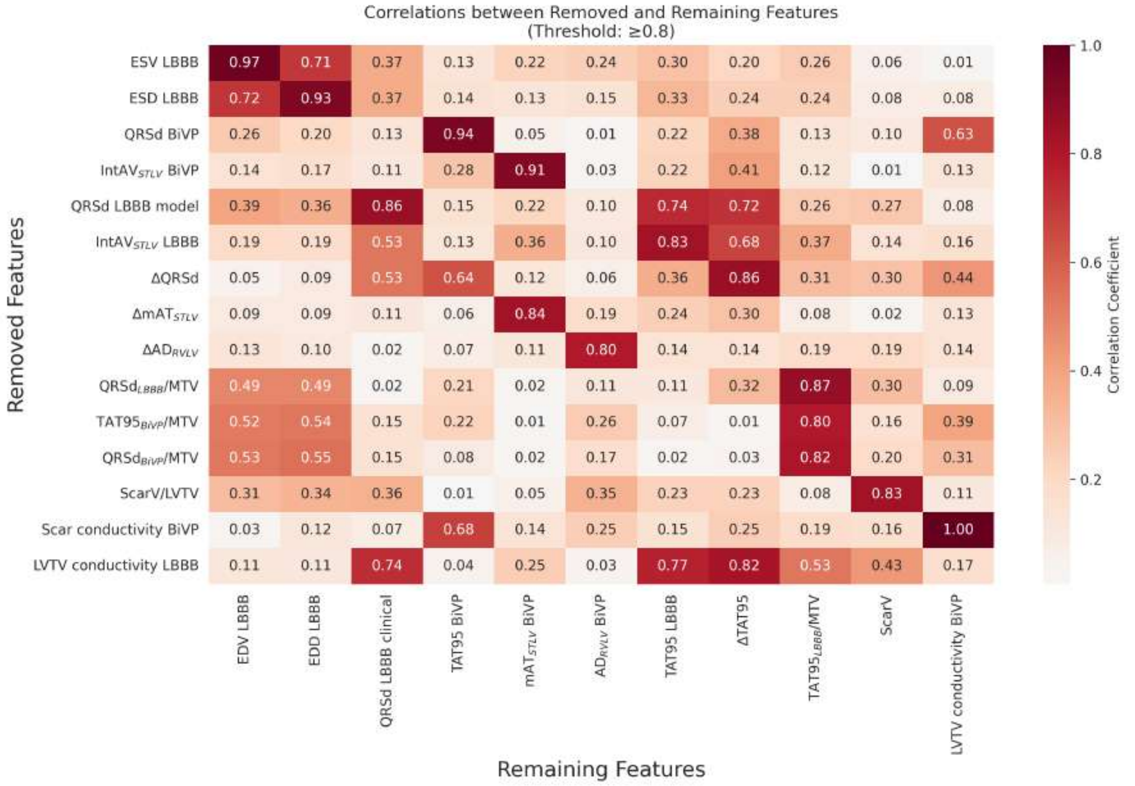


Figure S2: Pairwise correlations of removed features exceeding the collinearity threshold ($|r| \geq 0.8$) for the next training of the ML classifier.

$$AT_{RVLV} = ATmax_{LV} - ATmax_{RV}$$

$$mAT_{STLV} = \frac{LVlat_{mean} - ST_{mean}}{TAT}$$

$$IntAV_{STLV} = \int \left| \frac{AV_{LAT}(t)}{V_{LAT}} - \frac{AV_{ST}(t)}{V_{ST}} \right| dt$$

To quantify the improvement from the baseline LBBB to biventricular pacing, we calculated the relative change ($\Delta$) for each index as follows:

$$\Delta QRSd = \frac{\mathrm{QRSd}^{\mathrm{LBBB}} - \mathrm{QRSd}^{\mathrm{BiVP}}}{\mathrm{QRSd}^{\mathrm{LBBB}}}$$

$$\Delta\mathrm{TAT95} = \frac{\mathrm{TAT95}^{\mathrm{LBBB}} - \mathrm{TAT95}^{\mathrm{BiVP}}}{\mathrm{TAT95}^{\mathrm{LBBB}}}$$

$$\Delta\mathrm{AD}_{RVLV} = \frac{\mathrm{AD}_{RVLV}{}^{\mathrm{LBBB}} - \mathrm{AD}_{RVLV}{}^{\mathrm{BiVP}}}{\mathrm{AD}_{RVLV}{}^{\mathrm{LBBB}}}$$

$$\Delta\mathrm{mAT}_{STLV} = \frac{\mathrm{mAT}_{STLV}{}^{\mathrm{LBBB}} - \mathrm{mAT}_{STLV}{}^{\mathrm{BiVP}}}{\mathrm{mAT}_{STLV}{}^{\mathrm{LBBB}}}$$

$$\Delta\mathrm{IntAV}_{STLV} = \frac{\mathrm{IntAV}_{STLV}{}^{\mathrm{LBBB}} - \mathrm{IntAV}_{STLV}{}^{\mathrm{BiVP}}}{\mathrm{IntAV}_{STLV}{}^{\mathrm{LBBB}}}$$

where ST = septum (2th, 3th, 8th, 9th, 14th AHA segments); lat = LV lateral wall (5th, 6th, 11th, 12th, 16th AHA segments); AT = activation time; AV = activated volume; $V_{LAT}$ = a volume of the LV myocardium free wall, $V_{ST}$ = a volume of the interventricular septum myocardium.

| Variable Name, units | Description | Total group (n=74) | Responders (n=30) | Nonresponders (n=44) | Non-CS group (n=55) | CS group (n=19) |
|---|---|---|---|---|---|---|
| **Clinical-derived indices** | | | | | | |
| Age, year | Patient age at time of cardiac resynchronization therapy implantation | 63.6±6.6 | 64.0±6.1 | 63.5±7.0 | 63.1±6.1 | 65.4±7.9 |
| Gender (male/female) | Biological sex (male/female) | 52/22 | 21/9 | 31/13 | 37/18 | 15/4 |
| BMI, kg/m$^2$ | Body mass index calculated as weight in kilograms divided by height in meters squared | 28.1±4.7 | 26.8±5.1 | 29.0±4.2 | 28.3±4.6 | 27.7±5.0 |
| History of AF (1/0) | Binary indicator of atrial fibrillation history (1: present, 0: absent) | 11/63 | 4/26 | 10/34 | 10/45 | 1/18 |
| EF, % | Left ventricular ejection fraction measured baseline LBBB | 25.6±6.1 | 22.5±4.8 | 27.8±5.9 ** | 25.8±6.4 | 25.7±4.9 |
| QRSd LBBB, ms | QRS complex duration measured baseline LBBB | 189.2±22.8 | 192.0±20.4 | 187.3±24.3 | 191.7±23.7 | 182.0±18.6 # |
| EDV LBBB, ml | End-diastolic volume of left ventricle measured baseline LBBB | 281.2±82.0 | 291.4±76.7 | 274.4±86.3 | 291.9±82.2 | 250.7±77.6 # |
| ESV LBBB, ml | End-systolic volume of left ventricle measured baseline LBBB | 207.9±68.1 | 224.4±59.8 | 197.1±71.6 * | 216.0±69.1 | 185.1±61.2 |
| EDD LBBB, mm | Left ventricular end-diastolic diameter measured baseline LBBB | 72.7±7.4 | 73.0±8.3 | 72.4±7.0 | 73.8±7.4 | 69.5±7.0 # |
| ESD LBBB, mm | Left ventricular end-systolic diameter measured baseline LBBB | 62.4±8.6 | 63.4±9.0 | 61.7±8.4 | 63.2±8.8 | 59.8±7.9 |
| FC CHF (I/II/III) | Functional classification of congestive heart failure according to NYHA criteria | III: 59.5% (44)<br>II: 40.5% (30) | III: 50.0% (15)<br>II: 50.0% (15) | III: 65.9% (29)<br>II: 34.1% (15) | III: 60.0% (33)<br>II: 40.0% (22) | III: 57.9% (11)<br>II: 42.1% (8) |
| **CT/MRI derived indices** | | | | | | |
| Scar-LVPS distance, mm | Geodesic distance from LV pacing site to fibrosis region, computed using isotropic Eikonal equation | 31.4±25.5 | 40.36±25.5 | 25.5±23.9 * | 35.9±24.4 | 18.7±24.8 ## |
| RVPS-LVPS distance, mm | Geodesic distance between right and left ventricular pacing sites, computed using isotropic Eikonal equation | 103.3±22.0 | 105.2±20.5 | 102.0±23.0 | 100.6±22.1 | 110.8±20.1 |
| LAA-LVPS distance, mm | Geodesic distance from LV pacing site to latest activated area baseline LBBB, computed using isotropic Eikonal equation | 48.8±25.2 | 41.6±22.2 | 53.5±26.1 * | 48.9±25.5 | 48.5±25.0 |
| MTV, ml | Total myocardial tissue volume | 328.5±139.4 | 318.8±137.3 | 334.6±140.4 | 358.8±141.6 | 240.1±89.2 ## |
| LVTV, ml | Left ventricular myocardial tissue volume | 187.6±95.3 | 193.8±97.0 | 183.5±95.0 | 205.3±98.0 | 137.2±66.4 ## |
| ScarV, ml | Infarct scar/Fibrosis volume | 42.2±29.1 | 34.7±30.2 | 47.2±7.6 * | 42.8±30.7 | 40.8±24.6 |
| ScarV/LVTV, % | Percentage of fibrosis (Infacrt scar) tissue relative to LV myocardial tissue volume | 0.2±0.13 | 0.16±0.10 | 0.23±0.14 | 0.18±0.12 | 0.24±0.15 |
| **Model-derived indices** | | | | | | |
| AHA segment, № | Segment number from 17-segment American Heart Association model indicating left ventricular pacing site (LVPS) location during biventricular pacing (varied from 1 to 17) | 11: 28.4% (21)<br>12: 21.6% (16)<br>6: 20.3% (15)<br>5: 14.9% (11)<br>7: 9.5% (7)<br>16: 4.1% (3)<br>10: 1.4% (1) | 11: 30.0% (9)<br>12: 23.3% (7)<br>6: 23.3% (7)<br>5: 16.7% (5)<br>16: 6.7% (2) | 11: 27.3% (12)<br>12: 20.5% (9)<br>6: 18.2% (8)<br>7: 15.9% (7)<br>5: 13.6% (6)<br>10: 2.3% (1)<br>16: 2.3% (1) | 11: 32.7% (18)<br>5: 18.2% (10)<br>6: 18.2% (10)<br>12: 16.4% (9)<br>7: 9.1% (5)<br>16: 3.6% (2)<br>10: 1.8% (1) | 12: 36.8% (7)<br>6: 26.3% (5)<br>11: 15.8% (3)<br>7: 10.5% (2)<br>5: 5.3% (1)<br>16: 5.3% (1) |
| QRSd LBBB, ms | QRS duration from earliest onset to latest offset in LBBB | 190.5±21.6 | 192.6±20.7 | 189.1±22.2 | 191.2±22. 8 | 188.4±17.8 |
| TAT95 LBBB, ms | Total activation time (time for 95 % of myocardium to activate) in LBBB | 199.9±65.0 | 201.3±63.6 | 198.9±66.7 | 195.3±64.2 | 212.7±67.2 |
| $AD_{RVLV}$ LBBB, ms | Interventricular dyssynchrony index in LBBB | 90.2±36.2 | 85.6±34.4 | 93.3±37.4 | 83.6±34.9 | 109.2±33.9 ## |
| $mAT_{STLV}$ LBBB, dimensionless # | Mean LV lateral/septal delay time in LBBB | 0.33±0.07 | 0.34±0.07 | 0.33±0.06 | 0.32±0.06 | 0.35±0.08 |
| $IntAV_{STLV}$ LBBB, ms | Intraventricular dyssynchrony index (temporal activation imbalance between LV lateral wall and septum) in LBBB | 76.0±29.6 | 76.5±27.4 | 75.7±31.3 | 72.5±27.4 | 86.1±34.0 |
| QRSd BiVP, ms | QRS duration from earliest onset to latest offset in BiVP | 148.3±19.3 | 144.4±12.11 | 151.0±22.6 | 148.5±19.6 | 148.0±18.8 |
| TAT95 BiVP, ms | Total activation time (time for 95 % of myocardium to activate) in BiVP | 108.9±19.2 | 104.2±14.0 | 112.0±21.6 | 106.8±19.2 | 115.0±18.5 |
| $AD_{RVLV}$ BiVP, ms | Interventricular dyssynchrony index in BiVP | -0.7±20.9 | -5.2±19.5 | 2.30±21.5 | -4.6±19.1 | 10.4±22.3 # |
| $mAT_{STLV}$ BiVP, dimensionless | Mean LV lateral/septal delay time in BiVP | 0.14±0.08 | 0.14±0.08 | 0.14±0.07 | 0.15±0.08 | 0.11±0.06 |
| $IntAV_{STLV}$ BiVP, ms | Intraventricular dyssynchrony index (temporal activation imbalance between LV lateral wall and septum) in BiVP | 18.9±9.4 | 17.7±9.3 | 19.7±9.5 | 20.0±9.3 | 16.0±9.3 |
| ΔQRSd, % | Relative change in QRS complex duration | -0.21±0.14 | -0.24±0.09 | -0.19±0.16 | -0.21±0.15 | -0.21±0.11 |
| ΔTAT95, % | Relative change in total activation time | -0.41±0.21 | -0.44±0.15 | -0.38±0.24 | -0.4±0.23 | -0.42±0.15 |
| $\Delta AD_{RVLV}$, % | Relative change in interventricular dyssynchrony index | -1.06±0.37 | -1.12±0.29 | -1.03±0.41 | -1.12±0.39 | -0.91±0.22 # |
| $\Delta mAT_{STLV}$, % | Relative change in mean septal-to-lateral delay | -0.19±0.11 | -0.20±0.13 | -0.19±0.10 | -0.17±0.11 | -0.25±0.10 # |
| $\Delta IntAV_{STLV}$, % | Relative change in intraventricular dyssynchrony index | -57.1±33.7 | -58.9±32.2 | -55.9±34.9 | -52.5±32.2 | -70.1±35.1 |
| $QRSd_{LBBB}$/MTV, ms/ml | QRS duration during intrinsic LBBB normalized to total myocardial tissue volume | 0.67±0.26 | 0.71±0.29 | 0.65±0.24 | 0.61±0.24 | 0.85±0.23 ## |
| $TAT95_{LBBB}$/MTV, ms/ml | Total activation time during intrinsic LBBB normalized to total myocardial tissue volume | 0.71±0.37 | 0.76±0.47 | 0.67±0.28 | 0.61±0.31 | 0.96±0.41 ## |
| $QRSd_{BiVP}$/MTV, ms/ml | QRS duration during biventricular pacing normalized to total myocardial tissue volume | 0.52±0.19 | 0.53±0.22 | 0.51±0.17 | 0.47±0.17 | 0.67±0.16 ## |
| $TAT95_{BiVP}$/MTV, ms/ml | Total activation time during biventricular pacing normalized to total myocardial tissue volume | 0.38±0.15 | 0.39±0.17 | 0.38±0.14 | 0.33±0.12 | 0.52±0.15 ## |
| LVTV conductivity LBBB, mS/mm | Conductivity parameter assigned to viable myocardial tissue in the isotropic Eikonal equation during simulated LBBB | 0.18±0.12 | 0.16±0.12 | 0.19±0.13 | 0.19±0.14 | 0.15±0.09 |
| Scar conductivity LBBB, mS/mm | Conductivity parameter assigned to fibrosis tissue in the isotropic Eikonal equation during simulated LBBB | 0.06±0.04 | 0.05±0.04 | 0.06±0.04 | 0.06±0.05 | 0.05±0.03 |
| LVTV conductivity BiVP, mS/mm | Conductivity parameter assigned to viable myocardial tissue in the isotropic Eikonal equation during simulated BiVP | 0.38±0.13 | 0.38±0.12 | 0.38±0.14 | 0.40±0.14 | 0.33±0.10 # |
| Scar conductivity BiVP, mS/mm | Conductivity parameter assigned to fibrosis tissue in the isotropic Eikonal equation during simulated BiVP | 0.13±0.04 | 0.13±0.04 | 0.13±0.05 | 0.13±0.05 | 0.11±0.03 # |

Table S1: Comprehensive characterization of clinical-derived, imaging-derived, and model-derived parameters in the overall CRT cohort (n=74), stratified by therapeutic response and fibrosis characteristics. Data presented as mean ± standard deviation for continuous variables and frequency distributions for categorical variables. Non-CS group (n=55) represents patients with simplified fibrosis geometry, while CS group (n=19) comprises patients with extensive fibrosis who underwent personalized modeling with MRI-based fibrosis incorporation into CT-derived biventricular anatomy, including detailed coronary sinus segmentation. * - p<0.05, ** - p<0.01 Responders vs Non-responders. # - p<0.05, ## - p<0.01 Non-CS group vs CS group. Comparison between two independent groups was carried out using the Mann-Whitney test for quantitative data and Pearson's chi-square test for qualitative data.

| Variable | OR [95% CI] | P-value |
|---|---|---|
| Clinical Data | | |
| Age | 1.01 [0.94-1.09] | 0.757 |
| Gender | 0.98 [0.35-2.70] | 0.966 |
| BMI | 0.90 [0.80-1.00] | 0.053 |
| IHD/DCM | 0.51 [0.18-1.41] | 0.195 |
| History of AF | 0.81 [0.22-3.07] | 0.760 |
| EDV | 1.00 [1.00-1.01] | 0.391 |
| ESV | 1.01 [1.00-1.01] | 0.088 |
| EDD | 1.01 [0.95-1.08] | 0.721 |
| ESD | 1.02 [0.97-1.08] | 0.416 |
| **EF** | **0.84 [0.75-0.93]** | **<0.001** |
| FC CHF | 0.52 [0.20-1.34] | 0.173 |
| QRSd LBBB clinic | 1.01 [0.99-1.03] | 0.397 |
| **LV lead in Lateral Wall** | **10.88 [1.33-88.89]** | **0.026** |
| Model-Derived Data | | |
| **Scar-LVPS Distance** | **1.02 [1.00-1.04]** | **0.028** |
| RVPS-LVPS Distance | 1.01 [0.99-1.03] | 0.482 |
| **LAA-LVPS Distance** | **0.98 [0.96-1.00]** | **0.040** |
| ScarV | 0.99 [0.97-1.00] | 0.130 |
| ScarV/LVTV | 0.03 [0.00-1.51] | 0.080 |
| TAT95 LBBB | 1.00 [0.99-1.01] | 0.833 |
| QRSd LBBB | 1.01 [0.99-1.03] | 0.506 |
| AD STLV LBBB | 5.68 [0.00-7734.27] | 0.637 |
| AD RV LV LBBB | 1.00 [0.98-1.01] | 0.561 |
| TAT95 BiVP | 0.98 [0.96-1.01] | 0.142 |
| QRSd BiVP | 0.98 [0.96-1.01] | 0.215 |
| AD STLV BiVP | 0.16 [0.00-77.04] | 0.557 |
| AD RV LV BiVP | 0.98 [0.96-1.01] | 0.143 |
| delta TAT95 | 0.20 [0.02-2.57] | 0.219 |
| delta QRSd | 0.06 [0.00-3.04] | 0.157 |
| delta AD STLV | 0.25 [0.00-15.70] | 0.508 |
| delta AD RV LV | 0.51 [0.13-1.97] | 0.330 |

Table S2: Univariate analysis of the feature significance for CRT response prediction (n=74). OR - odds ratio, 95%CI(confidence interval) and P-value were calculated using univariable logistic regression.

| | Observed clinical outcome | Clinical LVPS | | | Optimal LVPS at LV segment centre | | | |
|---|---|---|---|---|---|---|---|---|
| Patient ID | Resp./ Nonresp. | AHA LV segment | ML score | Positive/ Negative | Concordant opt. segment for CS LVPS | AHA LV segment | ML score | Positive/ Negative |
| 01 | 1 | 12 | 0.75 | 1 | 1 | 6 | 0.76 | 1 |
| 02 | 1 | 6 | 0.98 | 1 | 0 | 5 | 0.99 | 1 |
| 05 | 1 | 12 | 0.06 | 0 | 0 | 11 | 0.06 | 0 |
| 09 | 1 | 6 | 0.73 | 1 | 0 | 12 | 0.83 | 1 |
| 22 | 1 | 12 | 0.33 | 0 | 0 | 5 | 0.50 | 0 |
| 33 | 1 | 16 | 0.55 | 1 | 1 | 16 | 0.56 | 1 |
| **03** | **0** | **7** | **0.30** | **0** | **0** | **6** | **0.74** | **1** |
| **04** | **0** | **6** | **0.30** | **0** | **0** | **6** | **0.58** | **1** |
| 07 | 0 | 6 | 0.33 | 0 | 1 | 12 | 0.42 | 0 |
| 10 | 0 | 6 | 0.30 | 0 | 0 | 5 | 0.31 | 0 |
| 14 | 0 | 12 | 0.38 | 0 | 0 | 6 | 0.35 | 0 |
| 15 | 0 | 5 | 0.26 | 0 | 1 | 12 | 0.44 | 0 |
| 16 | 0 | 11 | 0.77 | 1 | 0 | 6 | 0.90 | 1 |
| 17 | 0 | 12 | 0.13 | 0 | 0 | 5 | 0.30 | 0 |
| **20** | **0** | **11** | **0.26** | **0** | **0** | **5** | **0.62** | **1** |
| **21** | **0** | **12** | **0.45** | **0** | **0** | **11** | **0.54** | **1** |
| 23 | 0 | 12 | 0.30 | 0 | 0 | 12 | 0.33 | 0 |
| **24** | **0** | **7** | **0.32** | **0** | **0** | **6** | **0.50** | **1** |
| 30 | 0 | 11 | 0.14 | 0 | 0 | 12 | 0.32 | 0 |

Table S3: Classification of patients from the CS group at the clinical LVPS and the LVPS with the highest ML-score located at the centers of the AHA LV segments. Shown are: Responder/Nonresponder coded as 1/0, number of LV segment according to 17-segment AHA LV model with LVPS location, ML-score value depending on the LVPS location, Positive/Negative CRT response prediction coded as 1/0 depending on the ML-score above/below the threshold of 0.5; Concordant segment for the optimal LVPS at the centre of the 17-segmental AHA model and at the CS tributaries coded as 1/0. The bold rows highlight patients classified as true negative at the clinical LVPS, who were re-classified into positive for optimal LVPS.
