## Supplementary Data - SHAP for clinical LVPS for "Personalized planning of cardiac resynchronization therapy through integration of coronary sinus geometry, clinical data, digital twins, and machine learning: visualization, stratification, and optimization"

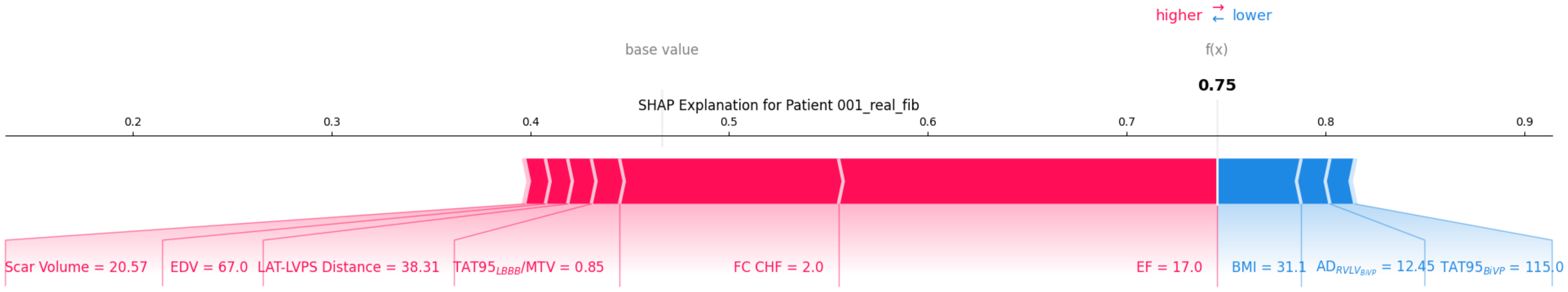

higher ⇄ lower
base value
f(x)
0.75
SHAP Explanation for Patient 001_real_fib
0.2
0.3
0.4
0.5
0.6
0.7
0.8
0.9
Scar Volume = 20.57
EDV = 67.0
LAT-LVPS Distance = 38.31
$TAT95_{LBBB}$/MTV = 0.85
FC CHF = 2.0
EF = 17.0
BMI = 31.1
$AD_{RVLV_{BiVP}}$ = 12.45
$TAT95_{BiVP}$ = 115.0

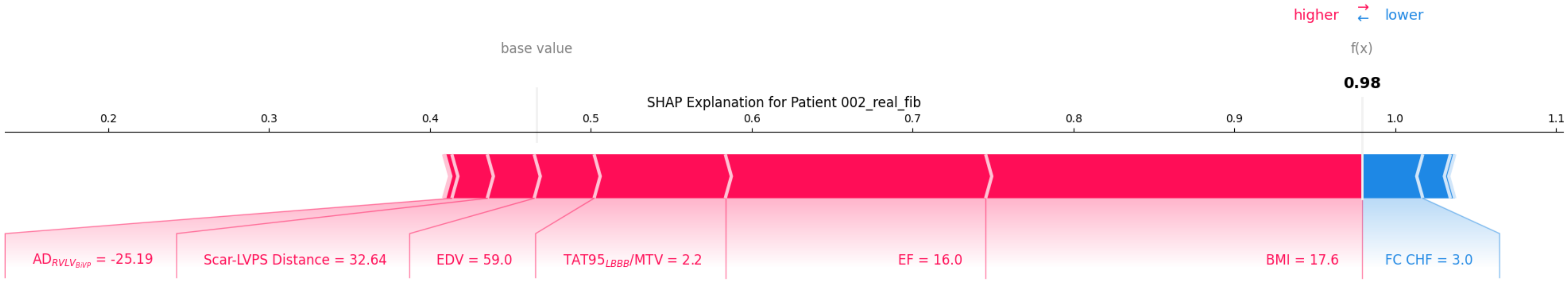

higher
lower
base value
f(x)
0.98
SHAP Explanation for Patient 002_real_fib
0.2
0.3
0.4
0.5
0.6
0.7
0.8
0.9
1.0
1.1
$AD_{RVLV_{BiVP}}$ = -25.19
Scar-LVPS Distance = 32.64
EDV = 59.0
$TAT95_{LBBB}$/MTV = 2.2
EF = 16.0
BMI = 17.6
FC CHF = 3.0

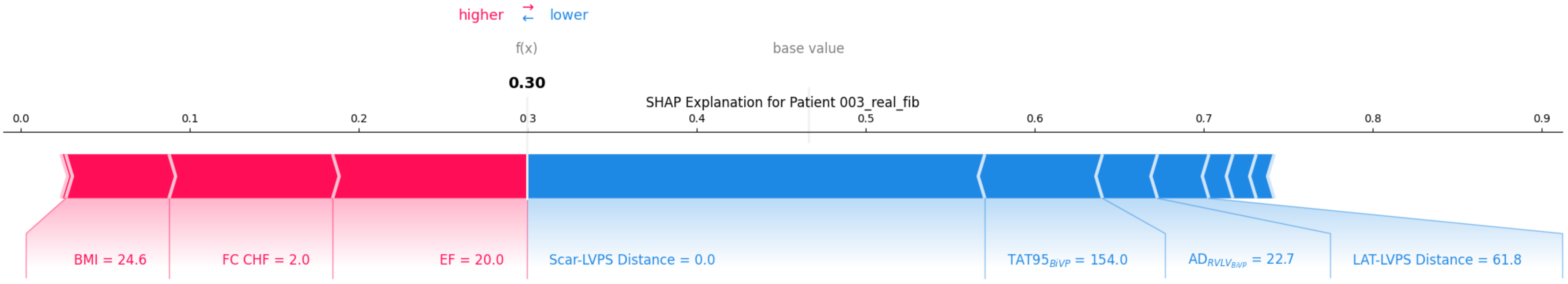

higher
lower
f(x)
base value
0.30
SHAP Explanation for Patient 003_real_fib
0.0
0.1
0.2
0.3
0.4
0.5
0.6
0.7
0.8
0.9
BMI = 24.6
FC CHF = 2.0
EF = 20.0
Scar-LVPS Distance = 0.0
$TAT95_{BiVP}$ = 154.0
$AD_{RVLV_{BiVP}}$ = 22.7
LAT-LVPS Distance = 61.8

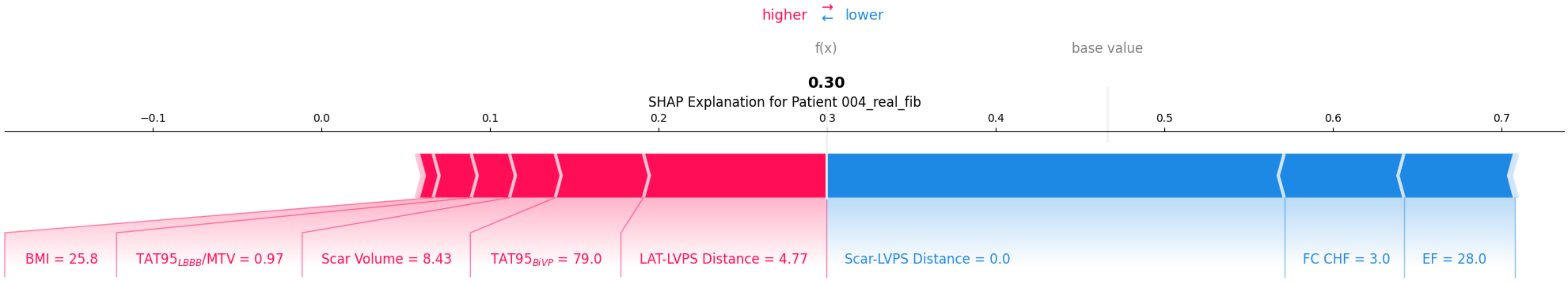

higher ⇄ lower
f(x)
base value
0.30
SHAP Explanation for Patient 004_real_fib
−0.1
0.0
0.1
0.2
0.3
0.4
0.5
0.6
0.7
BMI = 25.8
TAT95LBBB/MTV = 0.97
Scar Volume = 8.43
TAT95BiVP = 79.0
LAT-LVPS Distance = 4.77
Scar-LVPS Distance = 0.0
FC CHF = 3.0
EF = 28.0

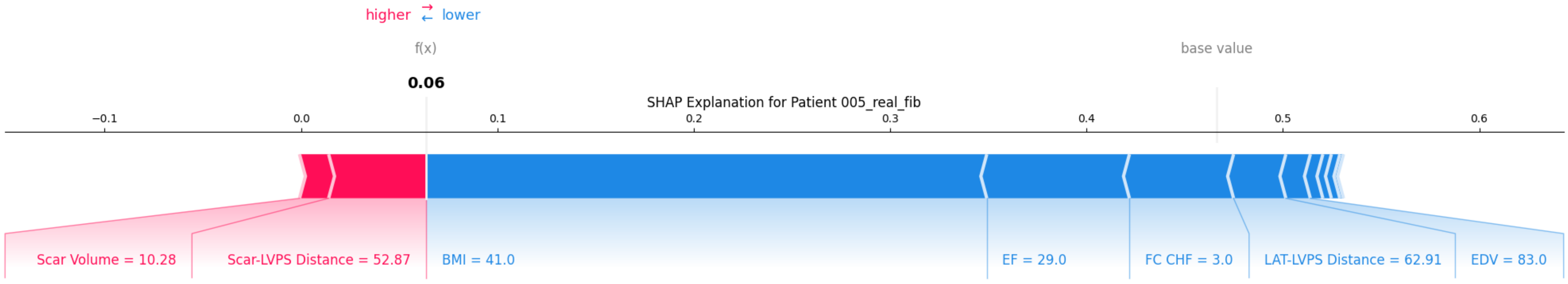
higher ⇄ lower
f(x)
0.06
base value
SHAP Explanation for Patient 005_real_fib
−0.1
0.0
0.1
0.2
0.3
0.4
0.5
0.6
Scar Volume = 10.28
Scar-LVPS Distance = 52.87
BMI = 41.0
EF = 29.0
FC CHF = 3.0
LAT-LVPS Distance = 62.91
EDV = 83.0

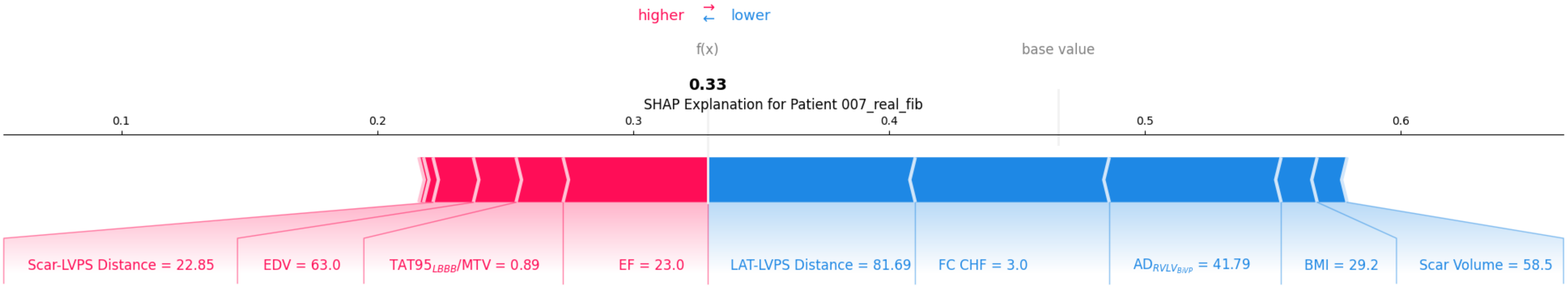
higher
lower
f(x)
base value
0.33
SHAP Explanation for Patient 007_real_fib
0.1
0.2
0.3
0.4
0.5
0.6
Scar-LVPS Distance = 22.85
EDV = 63.0
$TAT95_{LBBB}$/MTV = 0.89
EF = 23.0
LAT-LVPS Distance = 81.69
FC CHF = 3.0
$AD_{RVLV_{BiVP}}$ = 41.79
BMI = 29.2
Scar Volume = 58.5

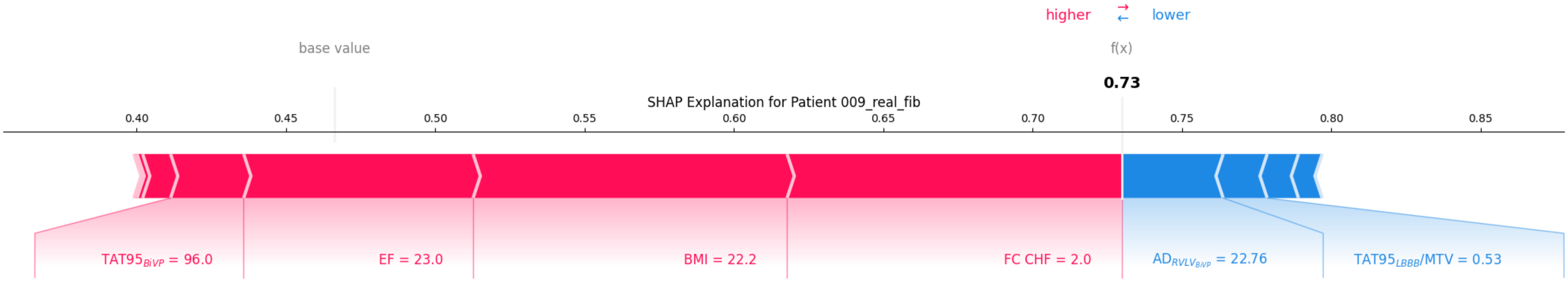
higher
lower
base value
f(x)
0.73
SHAP Explanation for Patient 009_real_fib
0.40
0.45
0.50
0.55
0.60
0.65
0.70
0.75
0.80
0.85
$TAT95_{BiVP}$ = 96.0
EF = 23.0
BMI = 22.2
FC CHF = 2.0
$AD_{RVLV_{BiVP}}$ = 22.76
$TAT95_{LBBB}$/MTV = 0.53

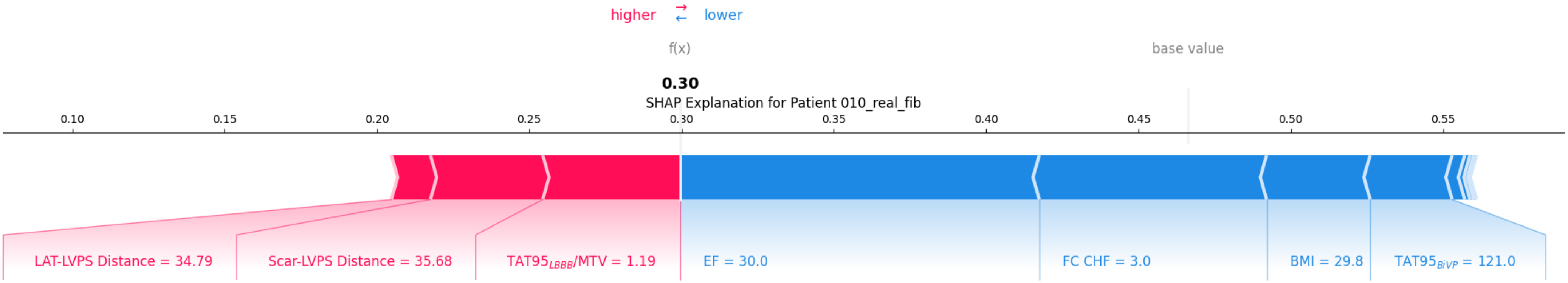
higher
lower
f(x)
base value
0.30
SHAP Explanation for Patient 010_real_fib
0.10
0.15
0.20
0.25
0.30
0.35
0.40
0.45
0.50
0.55
LAT-LVPS Distance = 34.79
Scar-LVPS Distance = 35.68
TAT95$_{LBBB}$/MTV = 1.19
EF = 30.0
FC CHF = 3.0
BMI = 29.8
TAT95$_{BiVP}$ = 121.0

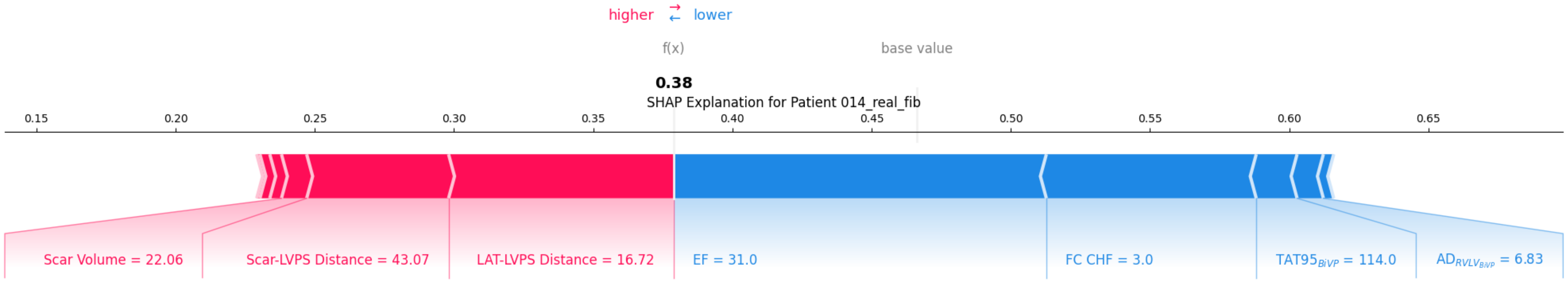
higher
lower
f(x)
base value
0.38
SHAP Explanation for Patient 014_real_fib
0.15
0.20
0.25
0.30
0.35
0.40
0.45
0.50
0.55
0.60
0.65
Scar Volume = 22.06
Scar-LVPS Distance = 43.07
LAT-LVPS Distance = 16.72
EF = 31.0
FC CHF = 3.0
TAT95$_{BiVP}$ = 114.0
AD$_{RVLV_{BiVP}}$ = 6.83

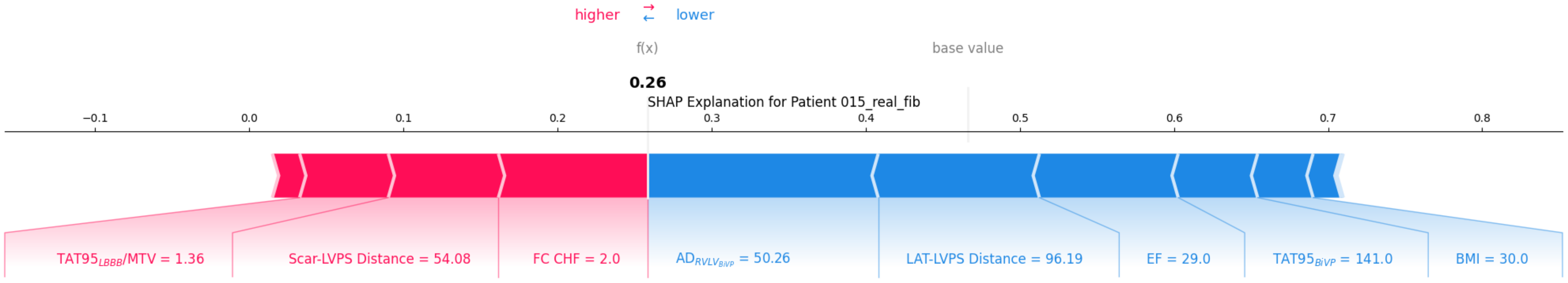
higher
lower
f(x)
base value
0.26
SHAP Explanation for Patient 015_real_fib
−0.1
0.0
0.1
0.2
0.3
0.4
0.5
0.6
0.7
0.8
TAT95$_{LBBB}$/MTV = 1.36
Scar-LVPS Distance = 54.08
FC CHF = 2.0
AD$_{RVLV_{BiVP}}$ = 50.26
LAT-LVPS Distance = 96.19
EF = 29.0
TAT95$_{BiVP}$ = 141.0
BMI = 30.0

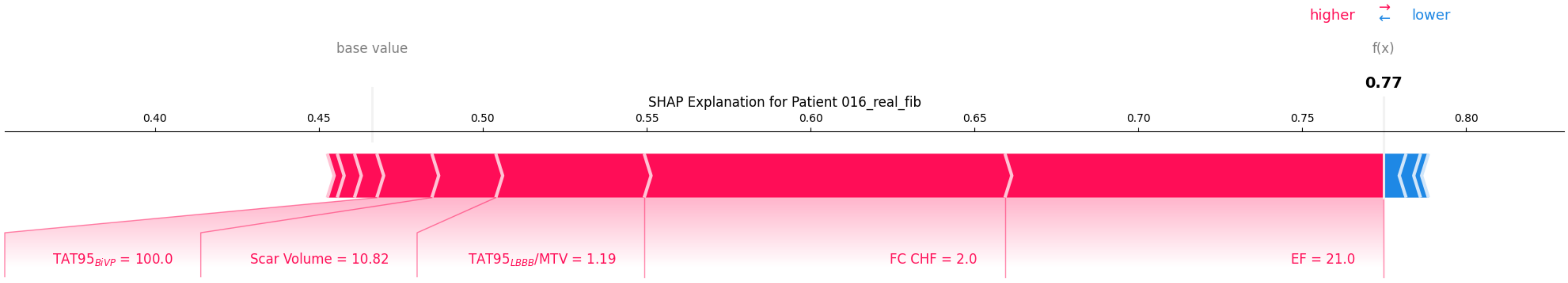
higher
lower
base value
f(x)
0.77
SHAP Explanation for Patient 016_real_fib
0.40
0.45
0.50
0.55
0.60
0.65
0.70
0.75
0.80
TAT95$_{BiVP}$ = 100.0
Scar Volume = 10.82
TAT95$_{LBBB}$/MTV = 1.19
FC CHF = 2.0
EF = 21.0

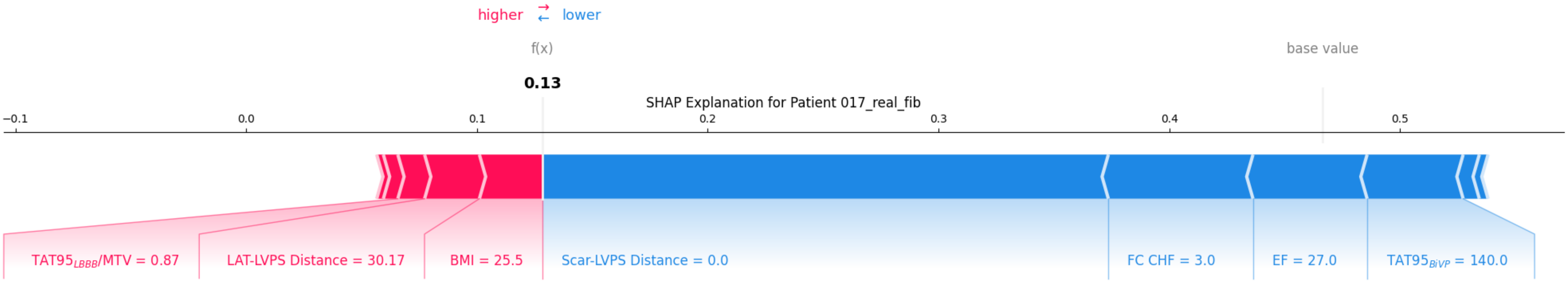

higher
lower
f(x)
base value
0.13
SHAP Explanation for Patient 017_real_fib
−0.1
0.0
0.1
0.2
0.3
0.4
0.5
TAT95$_{LBBB}$/MTV = 0.87
LAT-LVPS Distance = 30.17
BMI = 25.5
Scar-LVPS Distance = 0.0
FC CHF = 3.0
EF = 27.0
TAT95$_{BiVP}$ = 140.0

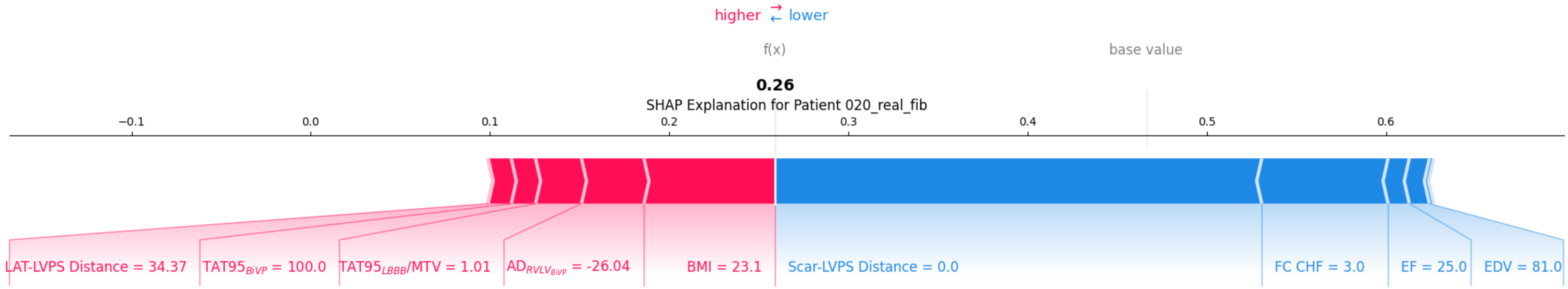

higher ⇄ lower
f(x)
base value
0.26
SHAP Explanation for Patient 020_real_fib
−0.1
0.0
0.1
0.2
0.3
0.4
0.5
0.6
LAT-LVPS Distance = 34.37
$TAT95_{BiVP}$ = 100.0
$TAT95_{LBBB}$/MTV = 1.01
$AD_{RVLV_{BiVP}}$ = -26.04
BMI = 23.1
Scar-LVPS Distance = 0.0
FC CHF = 3.0
EF = 25.0
EDV = 81.0

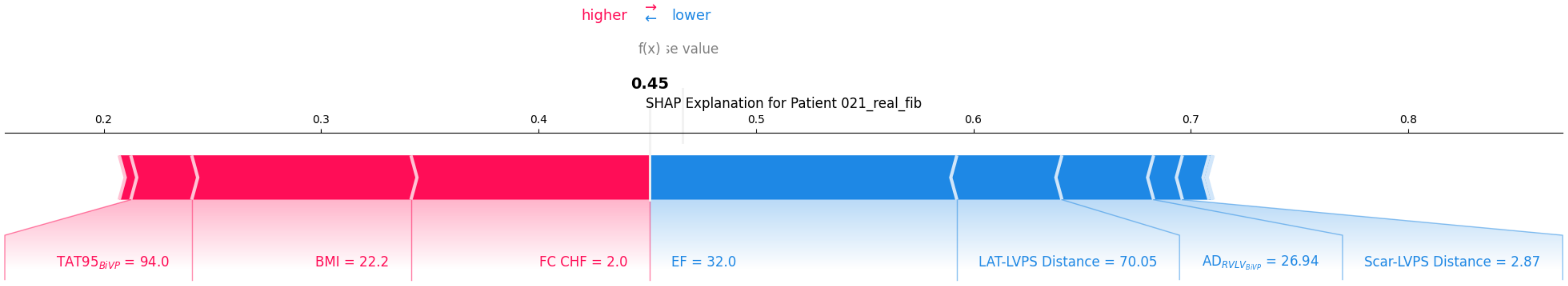

higher
lower
f(x)
se value
0.45
SHAP Explanation for Patient 021_real_fib
0.2
0.3
0.4
0.5
0.6
0.7
0.8
TAT95$_{BiVP}$ = 94.0
BMI = 22.2
FC CHF = 2.0
EF = 32.0
LAT-LVPS Distance = 70.05
AD$_{RVLV_{BiVP}}$ = 26.94
Scar-LVPS Distance = 2.87

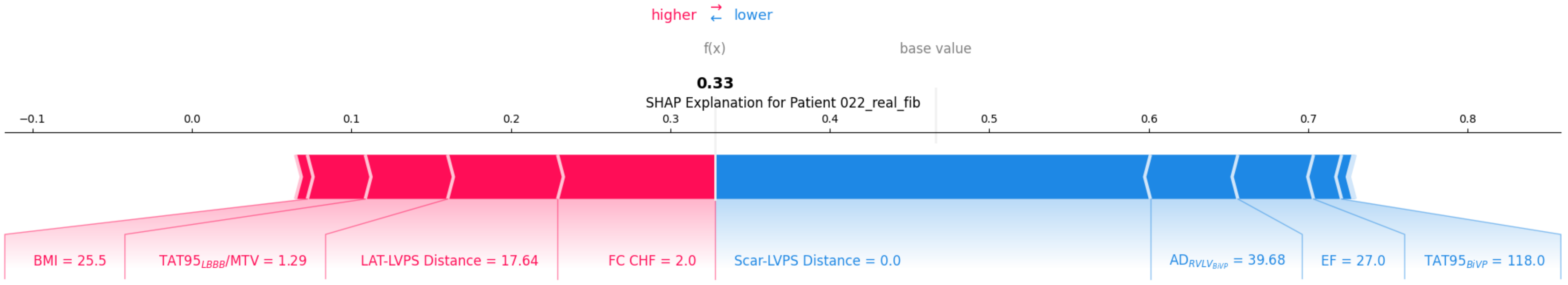
higher ⇄ lower
f(x)
base value
0.33
SHAP Explanation for Patient 022_real_fib
−0.1
0.0
0.1
0.2
0.3
0.4
0.5
0.6
0.7
0.8
BMI = 25.5
$TAT95_{LBBB}$/MTV = 1.29
LAT-LVPS Distance = 17.64
FC CHF = 2.0
Scar-LVPS Distance = 0.0
$AD_{RVLV_{BiVP}}$ = 39.68
EF = 27.0
$TAT95_{BiVP}$ = 118.0

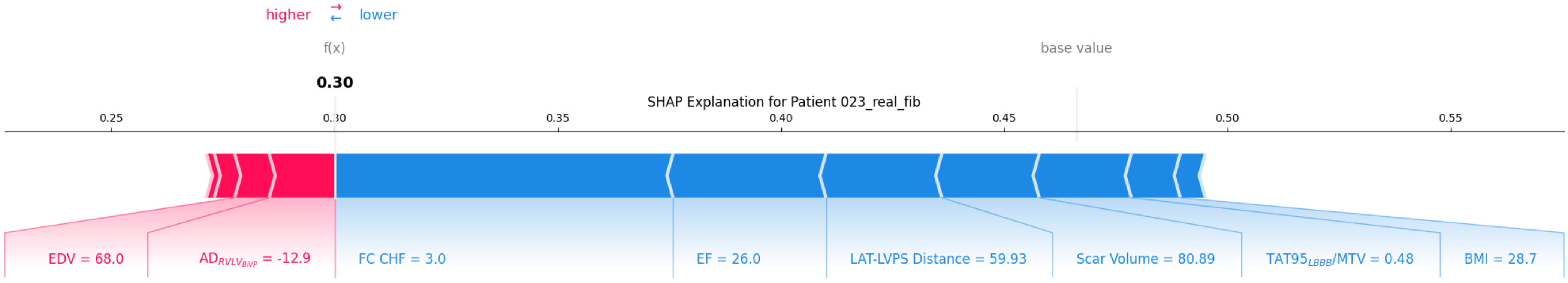

higher
lower
f(x)
base value
0.30
SHAP Explanation for Patient 023_real_fib
0.25
0.30
0.35
0.40
0.45
0.50
0.55
EDV = 68.0
AD$_{RVLV_{BiVP}}$ = -12.9
FC CHF = 3.0
EF = 26.0
LAT-LVPS Distance = 59.93
Scar Volume = 80.89
TAT95$_{LBBB}$/MTV = 0.48
BMI = 28.7

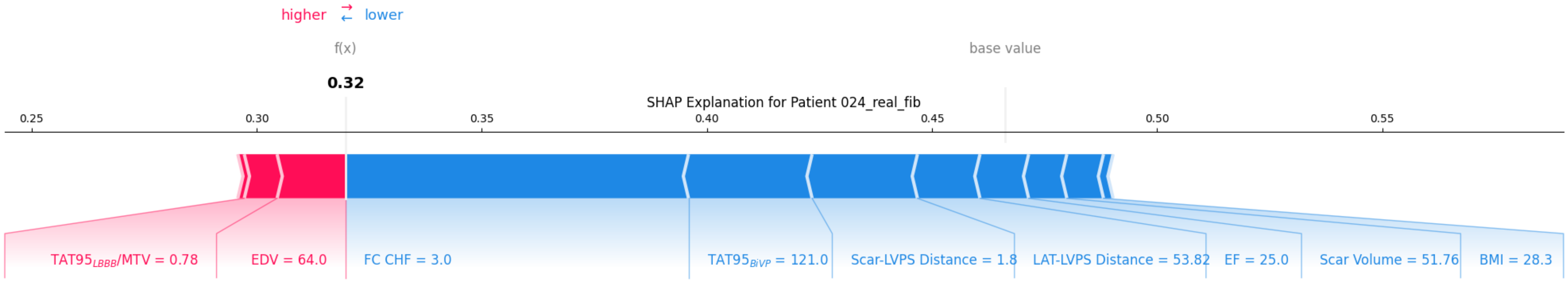
higher ⇄ lower
f(x)
base value
0.32
SHAP Explanation for Patient 024_real_fib
0.25
0.30
0.35
0.40
0.45
0.50
0.55
TAT95$_{LBBB}$/MTV = 0.78
EDV = 64.0
FC CHF = 3.0
TAT95$_{BiVP}$ = 121.0
Scar-LVPS Distance = 1.8
LAT-LVPS Distance = 53.82
EF = 25.0
Scar Volume = 51.76
BMI = 28.3

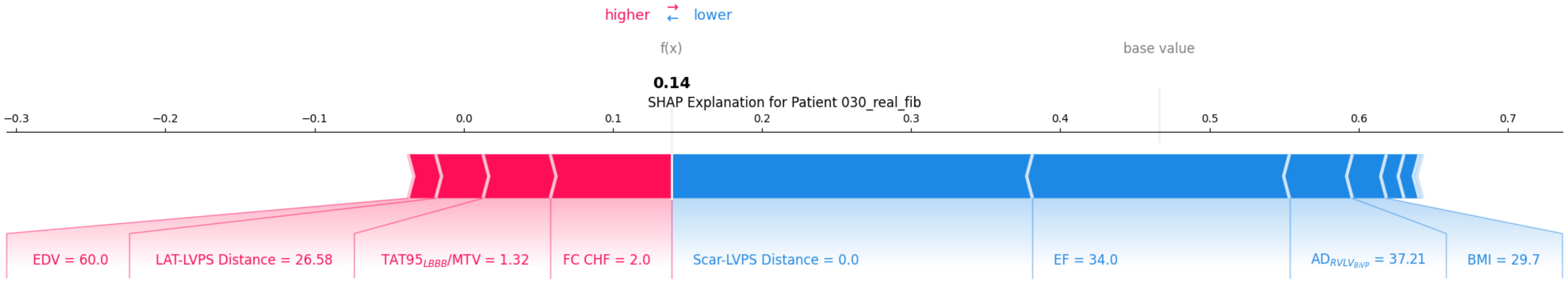
higher ⇄ lower
f(x)
base value
0.14
SHAP Explanation for Patient 030_real_fib
−0.3 −0.2 −0.1 0.0 0.1 0.2 0.3 0.4 0.5 0.6 0.7
EDV = 60.0
LAT-LVPS Distance = 26.58
TAT95$_{LBBB}$/MTV = 1.32
FC CHF = 2.0
Scar-LVPS Distance = 0.0
EF = 34.0
AD$_{RVLV_{BiVP}}$ = 37.21
BMI = 29.7

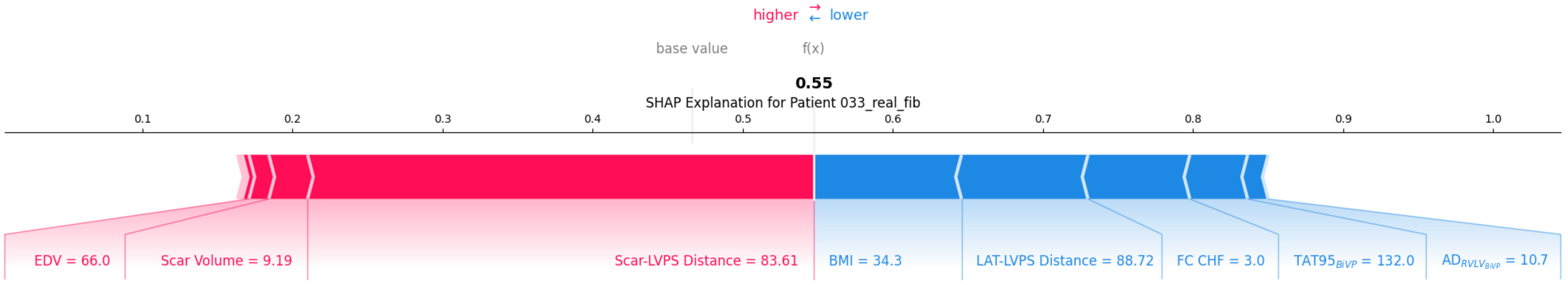
higher ⇄ lower
base value
f(x)
0.55
SHAP Explanation for Patient 033_real_fib
0.1
0.2
0.3
0.4
0.5
0.6
0.7
0.8
0.9
1.0
EDV = 66.0
Scar Volume = 9.19
Scar-LVPS Distance = 83.61
BMI = 34.3
LAT-LVPS Distance = 88.72
FC CHF = 3.0
$TAT95_{BiVP}$ = 132.0
$AD_{RVLV_{BiVP}}$ = 10.7
