## Supplementary Data - SHAP for optimal LVPS for "Personalized planning of cardiac resynchronization therapy through integration of coronary sinus geometry, clinical data, digital twins, and machine learning: visualization, stratification, and optimization"

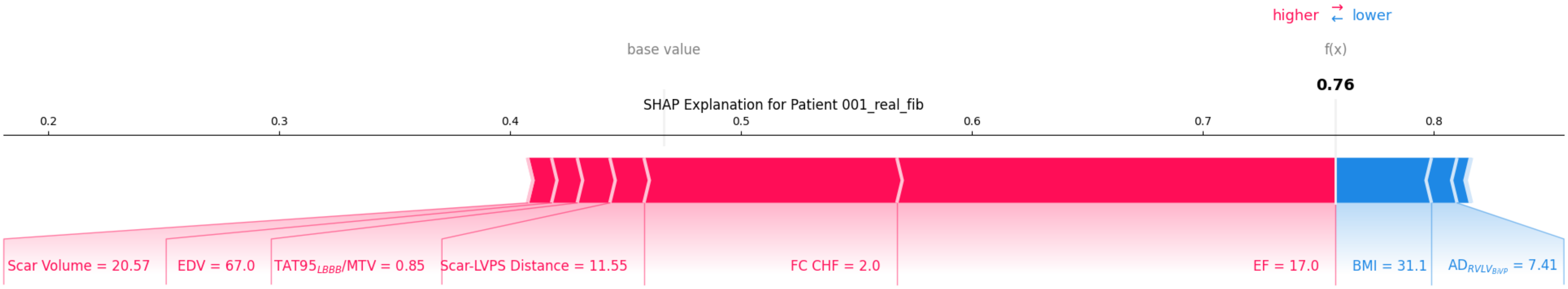

higher
lower
base value
f(x)
0.76
SHAP Explanation for Patient 001_real_fib
0.2
0.3
0.4
0.5
0.6
0.7
0.8
Scar Volume = 20.57
EDV = 67.0
TAT95$_{LBBB}$/MTV = 0.85
Scar-LVPS Distance = 11.55
FC CHF = 2.0
EF = 17.0
BMI = 31.1
AD$_{RVLV_{BiVP}}$ = 7.41

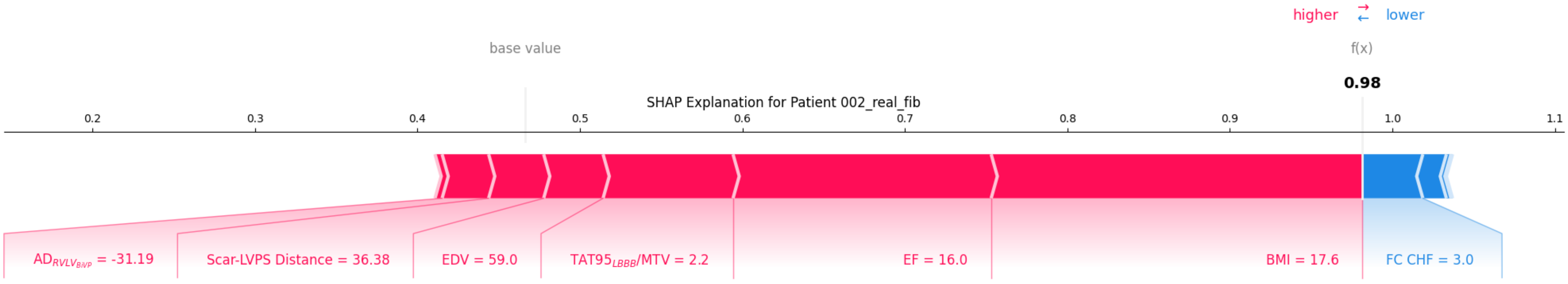
higher ⇄ lower
base value
f(x)
0.98
SHAP Explanation for Patient 002_real_fib
0.2
0.3
0.4
0.5
0.6
0.7
0.8
0.9
1.0
1.1
$AD_{RVLV_{BiVP}}$ = -31.19
Scar-LVPS Distance = 36.38
EDV = 59.0
$TAT95_{LBBB}$/MTV = 2.2
EF = 16.0
BMI = 17.6
FC CHF = 3.0

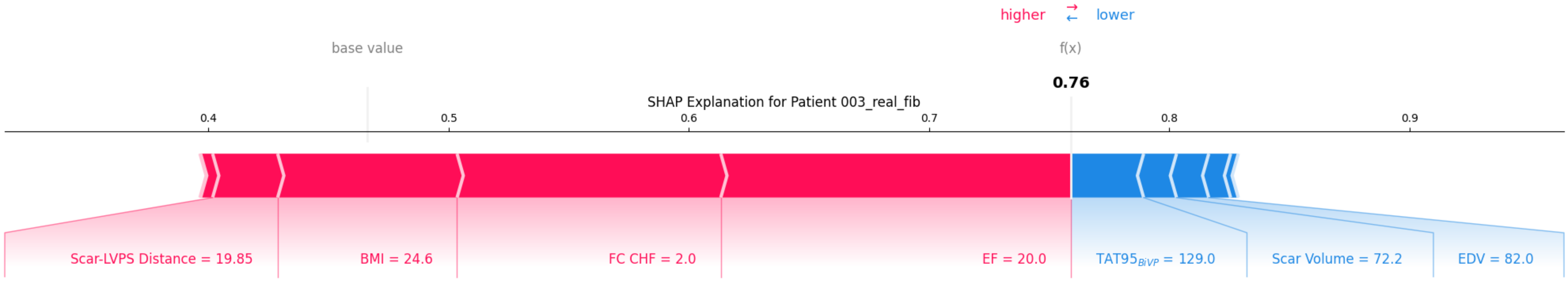

higher ⇄ lower
base value
f(x)
0.76
SHAP Explanation for Patient 003_real_fib
0.4
0.5
0.6
0.7
0.8
0.9
Scar-LVPS Distance = 19.85
BMI = 24.6
FC CHF = 2.0
EF = 20.0
$TAT95_{BiVP}$ = 129.0
Scar Volume = 72.2
EDV = 82.0

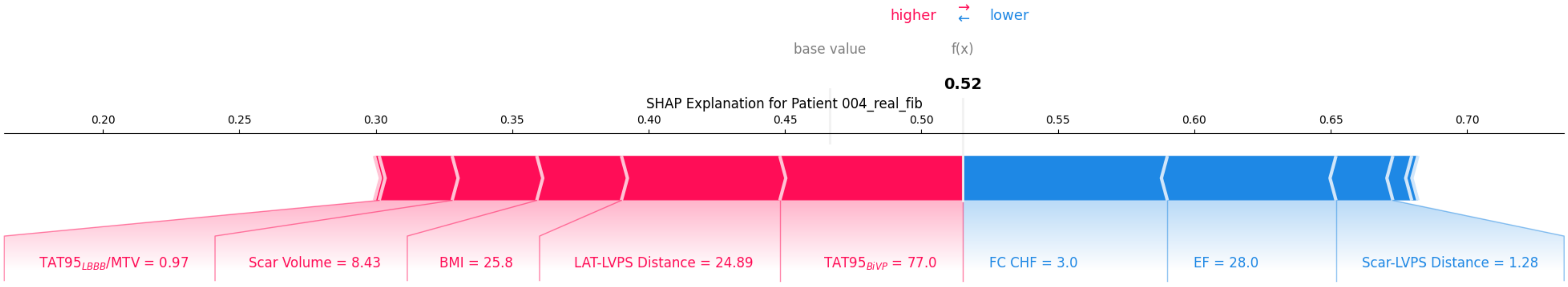
higher
lower
base value
f(x)
0.52
SHAP Explanation for Patient 004_real_fib
0.20
0.25
0.30
0.35
0.40
0.45
0.50
0.55
0.60
0.65
0.70
TAT95LBBB/MTV = 0.97
Scar Volume = 8.43
BMI = 25.8
LAT-LVPS Distance = 24.89
TAT95BiVP = 77.0
FC CHF = 3.0
EF = 28.0
Scar-LVPS Distance = 1.28

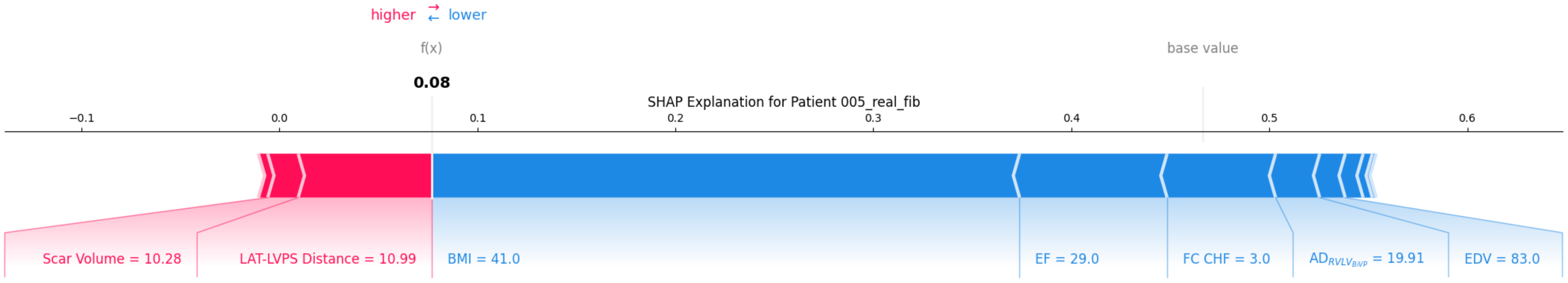

higher
lower
f(x)
base value
0.08
SHAP Explanation for Patient 005_real_fib
−0.1
0.0
0.1
0.2
0.3
0.4
0.5
0.6
Scar Volume = 10.28
LAT-LVPS Distance = 10.99
BMI = 41.0
EF = 29.0
FC CHF = 3.0
$AD_{RVLV_{BiVP}}$ = 19.91
EDV = 83.0

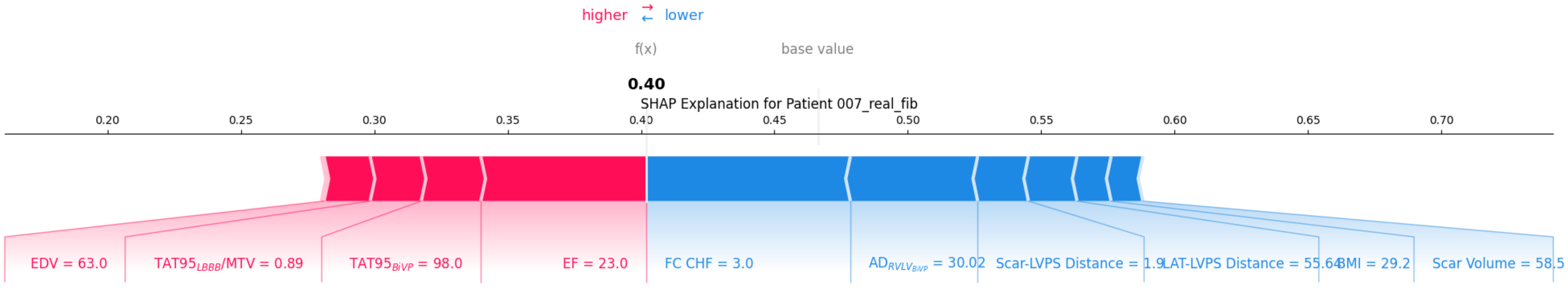
higher
lower
f(x)
base value
0.40
SHAP Explanation for Patient 007_real_fib
0.20
0.25
0.30
0.35
0.40
0.45
0.50
0.55
0.60
0.65
0.70
EDV = 63.0
TAT95LBBB/MTV = 0.89
TAT95BiVP = 98.0
EF = 23.0
FC CHF = 3.0
ADRVLVBiVP = 30.02
Scar-LVPS Distance = 1.9
LAT-LVPS Distance = 55.64
BMI = 29.2
Scar Volume = 58.5

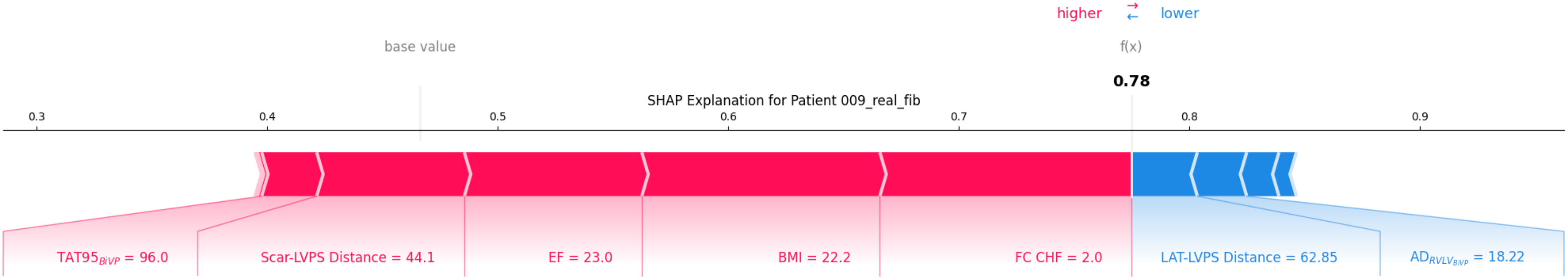
higher
lower
base value
f(x)
0.78
SHAP Explanation for Patient 009_real_fib
0.3
0.4
0.5
0.6
0.7
0.8
0.9
TAT95$_{BiVP}$ = 96.0
Scar-LVPS Distance = 44.1
EF = 23.0
BMI = 22.2
FC CHF = 2.0
LAT-LVPS Distance = 62.85
$AD_{RVLV_{BiVP}}$ = 18.22

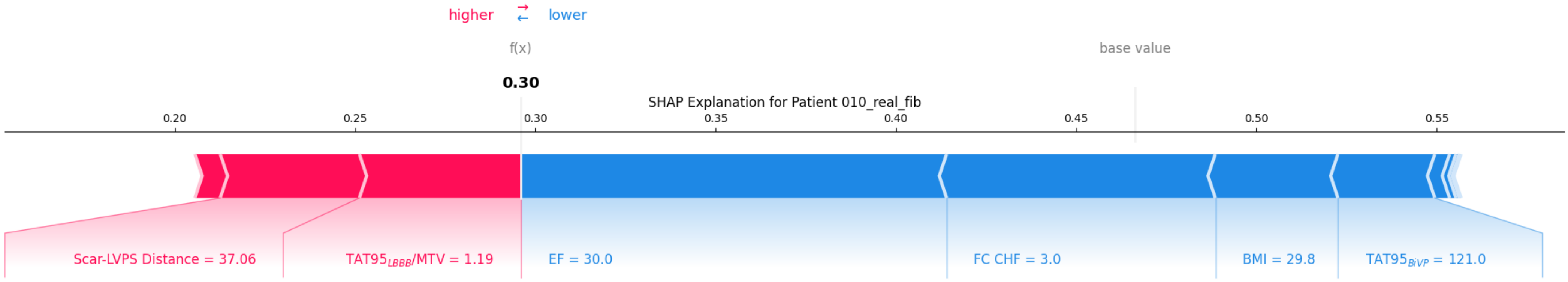
higher
lower
f(x)
base value
0.30
SHAP Explanation for Patient 010_real_fib
0.20
0.25
0.30
0.35
0.40
0.45
0.50
0.55
Scar-LVPS Distance = 37.06
TAT95LBBB/MTV = 1.19
EF = 30.0
FC CHF = 3.0
BMI = 29.8
TAT95BiVP = 121.0

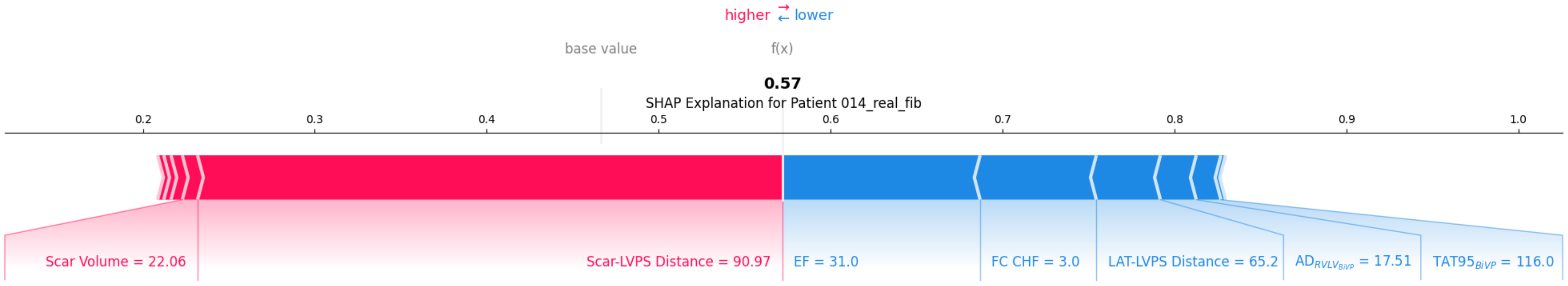

higher ⇄ lower
base value
f(x)
0.57
SHAP Explanation for Patient 014_real_fib
0.2
0.3
0.4
0.5
0.6
0.7
0.8
0.9
1.0
Scar Volume = 22.06
Scar-LVPS Distance = 90.97
EF = 31.0
FC CHF = 3.0
LAT-LVPS Distance = 65.2
$AD_{RVLV_{BiVP}}$ = 17.51
$TAT95_{BiVP}$ = 116.0

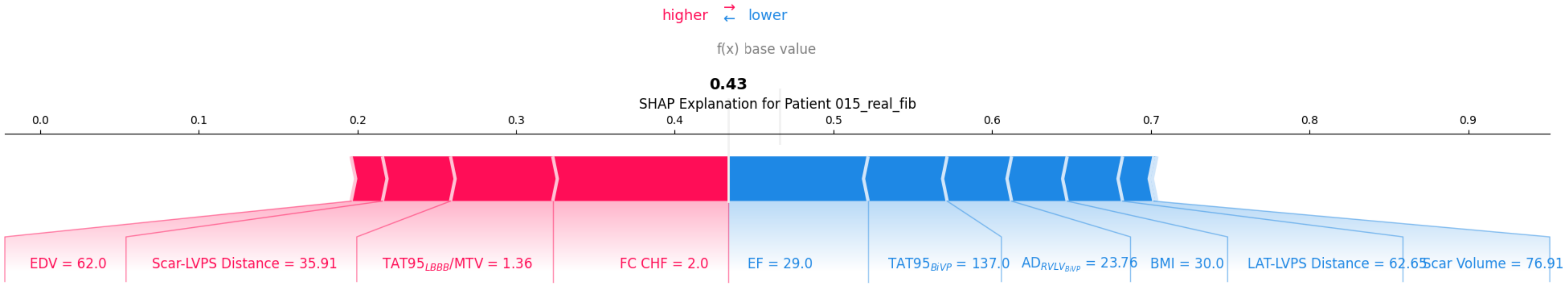

higher
lower
f(x)
base value
0.43
SHAP Explanation for Patient 015_real_fib
0.0
0.1
0.2
0.3
0.4
0.5
0.6
0.7
0.8
0.9
EDV = 62.0
Scar-LVPS Distance = 35.91
TAT95$_{LBBB}$/MTV = 1.36
FC CHF = 2.0
EF = 29.0
TAT95$_{BiVP}$ = 137.0
AD$_{RVLV_{BiVP}}$ = 23.76
BMI = 30.0
LAT-LVPS Distance = 62.65
Scar Volume = 76.91

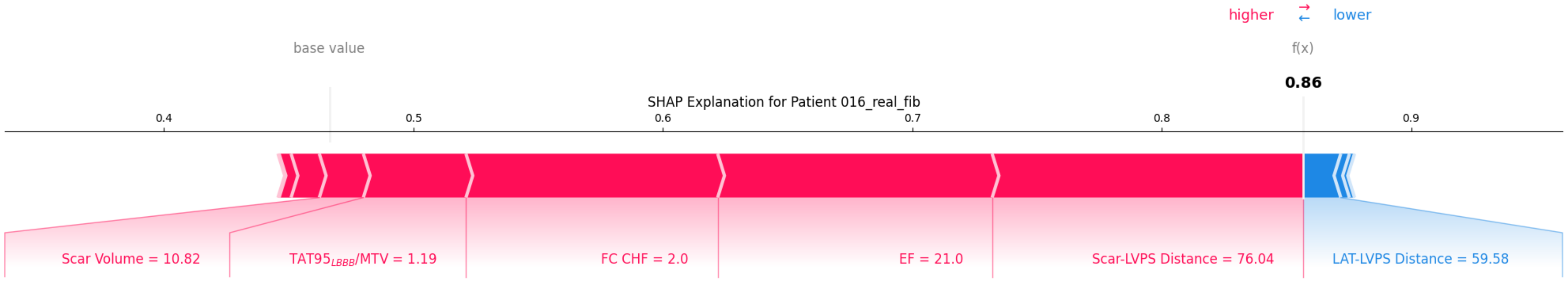

higher
lower
base value
f(x)
0.86
SHAP Explanation for Patient 016_real_fib
0.4
0.5
0.6
0.7
0.8
0.9
Scar Volume = 10.82
TAT95$_{LBBB}$/MTV = 1.19
FC CHF = 2.0
EF = 21.0
Scar-LVPS Distance = 76.04
LAT-LVPS Distance = 59.58

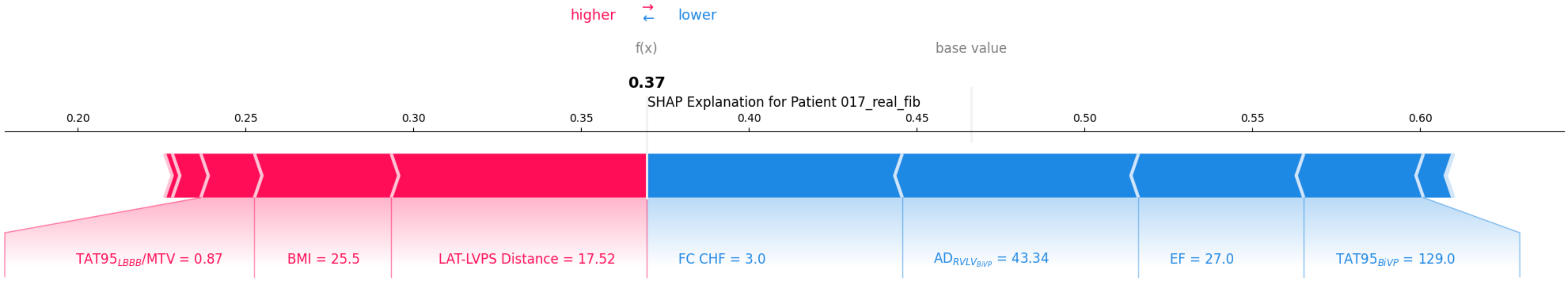
higher
lower
f(x)
base value
0.37
SHAP Explanation for Patient 017_real_fib
0.20
0.25
0.30
0.35
0.40
0.45
0.50
0.55
0.60
TAT95$_{LBBB}$/MTV = 0.87
BMI = 25.5
LAT-LVPS Distance = 17.52
FC CHF = 3.0
AD$_{RVLV_{BiVP}}$ = 43.34
EF = 27.0
TAT95$_{BiVP}$ = 129.0

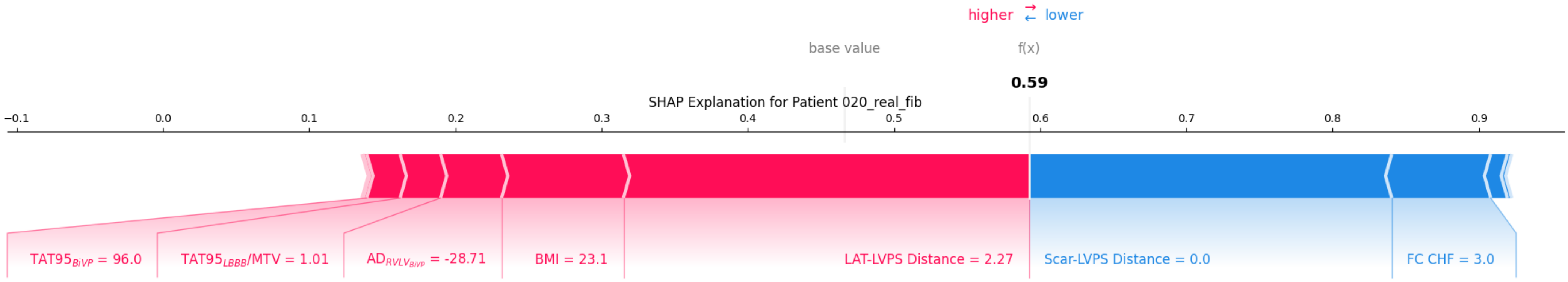
higher ⇄ lower
base value
f(x)
0.59
SHAP Explanation for Patient 020_real_fib
−0.1
0.0
0.1
0.2
0.3
0.4
0.5
0.6
0.7
0.8
0.9
TAT95$_{BiVP}$ = 96.0
TAT95$_{LBBB}$/MTV = 1.01
AD$_{RVLV_{BiVP}}$ = -28.71
BMI = 23.1
LAT-LVPS Distance = 2.27
Scar-LVPS Distance = 0.0
FC CHF = 3.0

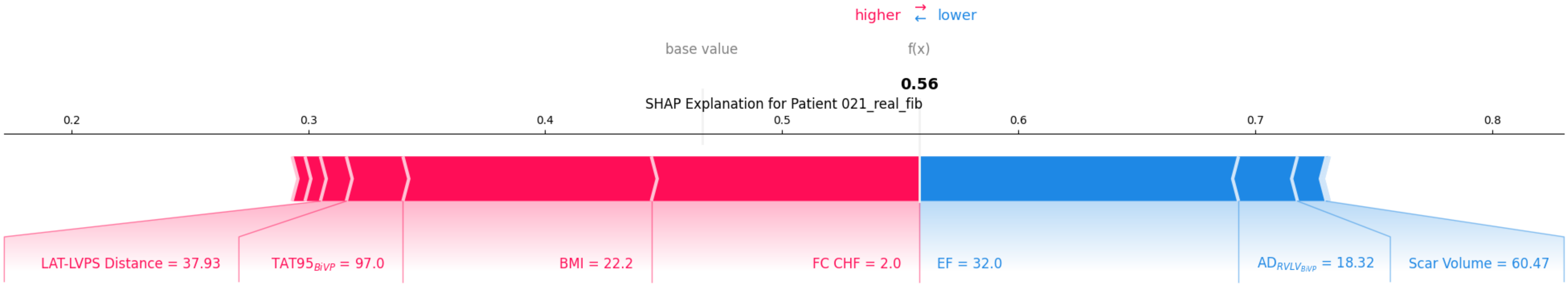

higher ⇄ lower
base value
f(x)
0.56
SHAP Explanation for Patient 021_real_fib
0.2
0.3
0.4
0.5
0.6
0.7
0.8
LAT-LVPS Distance = 37.93
$TAT95_{BiVP}$ = 97.0
BMI = 22.2
FC CHF = 2.0
EF = 32.0
$AD_{RVLV_{BiVP}}$ = 18.32
Scar Volume = 60.47

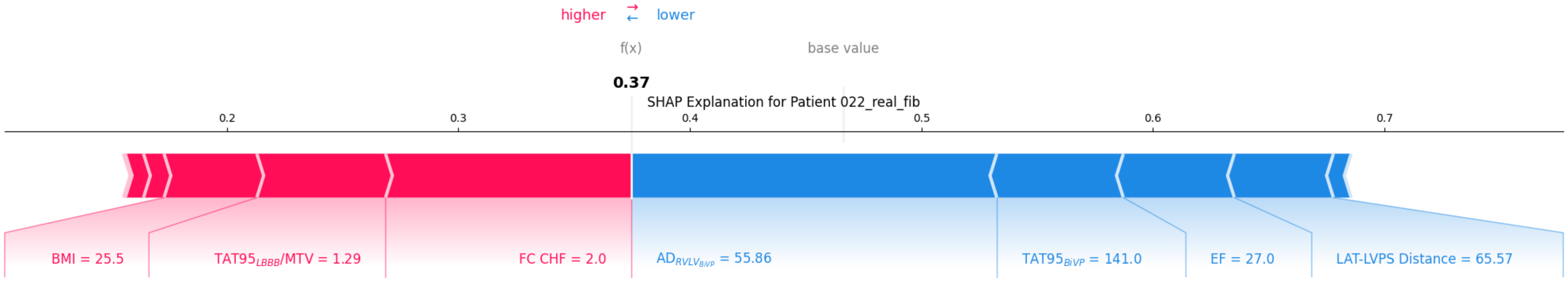
higher ⇄ lower
f(x)
base value
0.37
SHAP Explanation for Patient 022_real_fib
0.2
0.3
0.4
0.5
0.6
0.7
BMI = 25.5
$TAT95_{LBBB}$/MTV = 1.29
FC CHF = 2.0
$AD_{RVLV_{BiVP}}$ = 55.86
$TAT95_{BiVP}$ = 141.0
EF = 27.0
LAT-LVPS Distance = 65.57

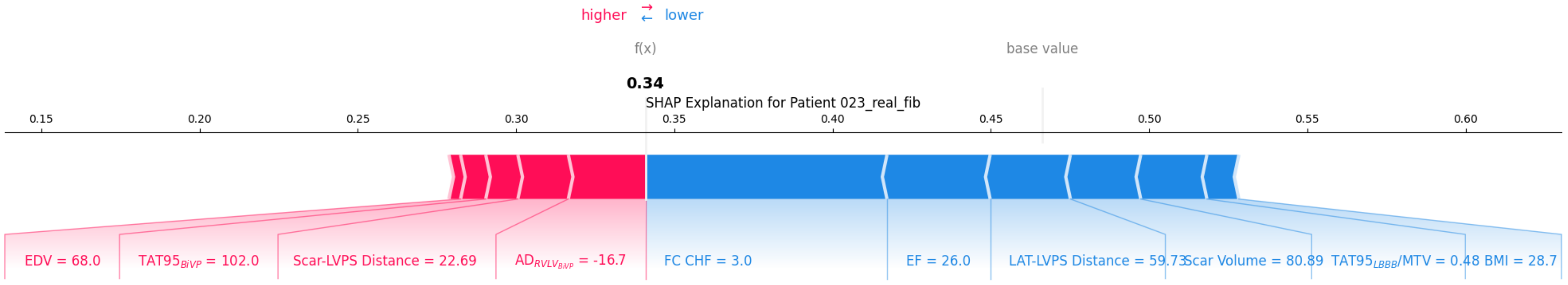
higher
lower
f(x)
base value
0.34
SHAP Explanation for Patient 023_real_fib
0.15
0.20
0.25
0.30
0.35
0.40
0.45
0.50
0.55
0.60
EDV = 68.0
$TAT95_{BiVP}$ = 102.0
Scar-LVPS Distance = 22.69
$AD_{RVLV_{BiVP}}$ = -16.7
FC CHF = 3.0
EF = 26.0
LAT-LVPS Distance = 59.73
Scar Volume = 80.89
$TAT95_{LBBB}$/MTV = 0.48
BMI = 28.7

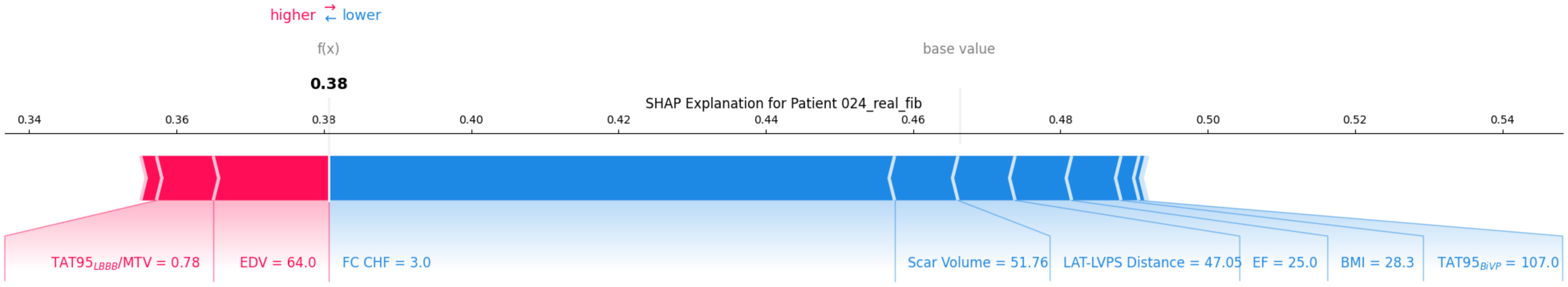
higher ⇄ lower
f(x)
base value
0.38
SHAP Explanation for Patient 024_real_fib
0.34
0.36
0.38
0.40
0.42
0.44
0.46
0.48
0.50
0.52
0.54
$TAT95_{LBBB}$/MTV = 0.78
EDV = 64.0
FC CHF = 3.0
Scar Volume = 51.76
LAT-LVPS Distance = 47.05
EF = 25.0
BMI = 28.3
$TAT95_{BiVP}$ = 107.0

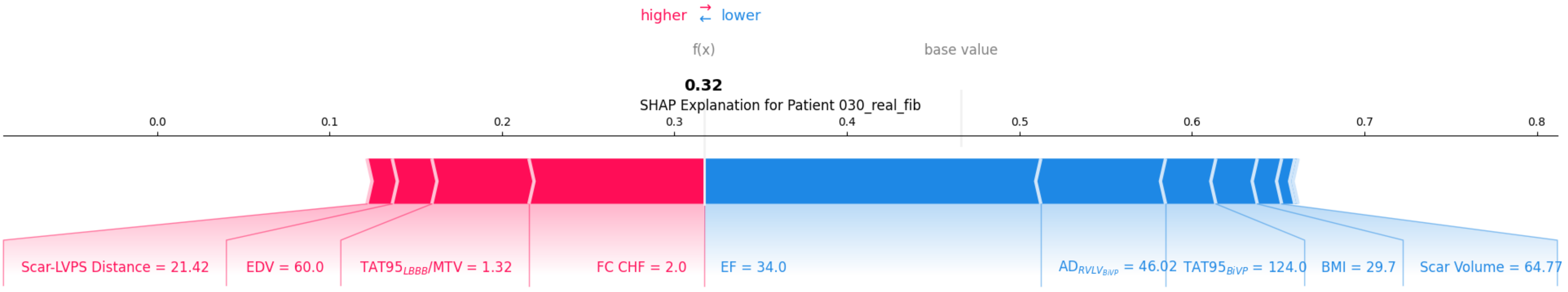
higher
lower
f(x)
base value
0.32
SHAP Explanation for Patient 030_real_fib
0.0
0.1
0.2
0.3
0.4
0.5
0.6
0.7
0.8
Scar-LVPS Distance = 21.42
EDV = 60.0
$TAT95_{LBBB}$/MTV = 1.32
FC CHF = 2.0
EF = 34.0
$AD_{RVLV_{BiVP}}$ = 46.02
$TAT95_{BiVP}$ = 124.0
BMI = 29.7
Scar Volume = 64.77

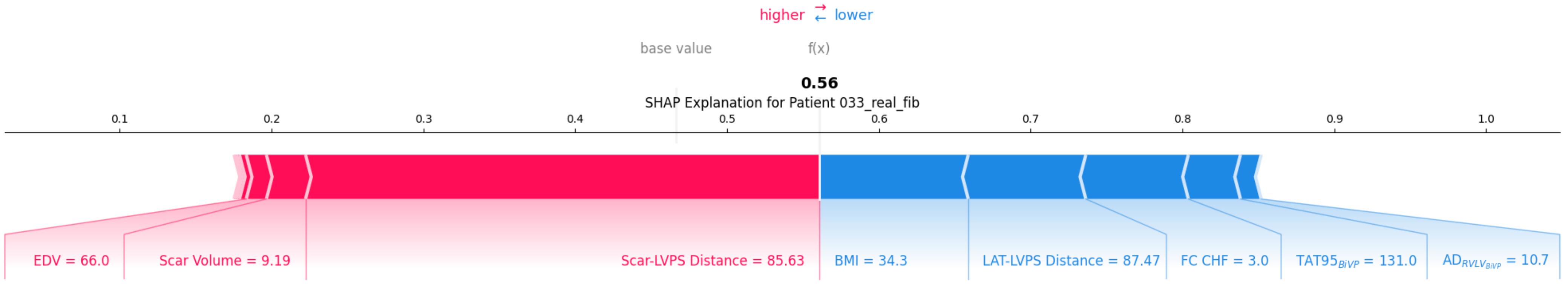

higher ⇄ lower
base value
f(x)
0.56
SHAP Explanation for Patient 033_real_fib
0.1
0.2
0.3
0.4
0.5
0.6
0.7
0.8
0.9
1.0
EDV = 66.0
Scar Volume = 9.19
Scar-LVPS Distance = 85.63
BMI = 34.3
LAT-LVPS Distance = 87.47
FC CHF = 3.0
$TAT95_{BiVP}$ = 131.0
$AD_{RVLV_{BiVP}}$ = 10.7
